# Defining and measuring unmet palliative care needs among people with life-limiting illness: a scoping review of international evidence

**DOI:** 10.1101/2025.07.07.25330993

**Authors:** Therese Johansson, Melanie Diggle, Anne Finucance, Joanna M Davies, Irene J Higginson, Katherine E Sleeman, Lorna K Fraser, Fliss E M Murtagh, Anna E Bone

## Abstract

**Background:** Quantifying palliative care needs and whether they are met is essential for effective service planning and provision. Estimates of palliative care needs are widely reported but less is known about unmet needs, with no accepted definition of this construct or guidance on how to measure it.

**Aim:** To investigate how unmet palliative care needs for adults with advanced life-limiting illness have been defined, measured, and reported in the evidence.

**Design:** Scoping review following the Joanna Briggs Institute guidelines.

**Data sources:** In October 2024, we systematically searched MEDLINE, EMBASE, CINAHL, and PsycINFO for quantitative and mixed-methods studies published after 2000. Additional backward and forward citation and manual searching of grey literature sources were performed. Data on the definitions, measurement and reporting of unmet needs were extracted and charted and summarised narratively using inductive content analysis and framework synthesis.

**Results:** Seventy studies were included: nine reviews and 61 primary evidence studies from 16 countries. Only 11 (16%) studies explicitly defined unmet palliative care needs, usually as a discrepancy between required and received care. We identified three approaches to measurement: of unmet palliative care needs: 1) *symptoms and concerns*; 2) *access to services*; and 3) *sufficiency of service provision to resolve symptoms and concerns*.

**Conclusions:** This novel review highlights a lack of consensus regarding the definition, measurement and reporting of unmet palliative care needs. Three approaches identified in the review emphasise different aspects of unmet needs: their strengths and limitations are discussed and guidance on their use is provided for various end-users.

**What is already known about the topic?:**

- Prevalence of palliative care needs are widely reported in research and policy, but how unmet needs are defined and measured is less well understood.

**What this paper adds:**

- Few studies provide a clear, detailed definition of unmet palliative care needs.
- We identify three main approaches to measuring unmet needs in palliative care research, by quantifying: 1) Symptoms and concerns; 2) Access to services; and 3) Sufficiency of service provision to resolve symptoms and concerns.
- There is little focus on non-cancer populations and few reports of involvement of patients and carers in studies measuring unmet palliative care needs.

**Implications for practice, theory or policy:**

- Methodological strengths and limitations of the three identified approaches to measuring unmet palliative care needs are discussed.
- To address the knowledge gaps identified, recommendations for reporting of definitions and how these are operationalised are provided.

## Background

With population ageing, rising multimorbidity, and increasing health-related suffering (1–4), providing timely and adequate palliative care is becoming a major public health priority. Despite palliative care being recognized as a human right (5), barriers to access persist, leaving some needs unmet. Unmet care needs are concerning at the individual, group and societal level. Assessing and quantifying unmet palliative care needs of a population is essential to inform effective planning and delivery of healthcare services and reduce inequitable access (6–8), yet guidance on how best to define and measure this construct is lacking.

In healthcare research, care needs are commonly defined using Bradshaw’s typology as the ‘ability to benefit from healthcare services’ (9). These needs can be further categorised as: 1) *felt need* (individual perceptions of need); 2) *expressed need* (perceived need resulting in demanded care); 3) *normative need* (defined by experts and thus largely based on healthcare professionals’ perceptions) and 4) *comparative need* (needs compared across different groups) (9). While other frameworks for conceptualising need exist, e.g., Maslow’s hierarchy of need, Bradshaw’s typology is particularly useful in palliative care as it highlights who determines the need (which influences what is reported as need) rather than simply the what the need is (10).

A needs assessment is a process to determine need for care services in relation to health problems, either at the individual level (e.g., clinical assessments of patients’ healthcare needs) or population-level (e.g., evaluating community needs for care services) (11). A review by Franks et al. (12) identified two main methods for population-level palliative care needs assessment: an epidemiological approach based on symptom prevalence among patients in advanced stages of life-limiting conditions presumed to benefit from palliative care; and an approach based on healthcare service use. It is also vital to understand how many do not have their palliative care needs met, but the lack of an accepted definition of unmet palliative care needs makes estimations difficult.

The present review expands on Franks et al.’s findings by focusing on *unmet* palliative care needs while broadening the scope beyond prior reviews, which have often focused on unmet needs in specific patient groups or care settings(13, 14).To inform our search strategies for, we constructed operational definitions of ‘palliative care need’ and ‘unmet palliative care need’ from definitions of needs and unmet needs in the broader health and care literature (15–18) and the World Health Organisation’s classification of palliative care as a holistic care approach to improve the quality of life of patients with life-limiting illness and their families (5, 19). To operationalise this, we defined palliative care needs as “the ability for people with life-limiting illness to benefit from available care to manage a range of symptoms and concerns including physical, psychological, social, spiritual, informational, and practical”. These needs can be addressed by specialist and/or generalist palliative care professionals, or a combination of both (20). We defined unmet palliative care needs as “the difference or gap between required or expected care and actual care received by people with life-limiting illness”.

This review aimed to clarify concepts in the literature by addressing the question: How have unmet palliative care needs for adults with advanced life-limiting illness been defined, measured, and reported in the evidence?

## Methods

### Design

This scoping review follows the methodological guidelines set out by the Joanna Briggs Institute (JBI) (21) and is reported according to the Preferred Reporting Items for Systematic Reviews and Meta Analyses extension for Scoping Reviews (PRISMA ScR) (22). The review protocol was registered on the Open Science Framework (registration reference: https://doi.org/10.17605/OSF.IO/M8DHA).

### Eligibility criteria

We included studies reporting quantitative data on how unmet needs of adults (aged 18+ years) with advanced life-limiting illness were defined, measured and reported. Table 1 presents the full inclusion and exclusion criteria. Peer-reviewed quantitative and mixed-methods studies were included, as were systematic and scoping reviews reporting results from quantitative and mixed-methods studies. Grey literature (reports but not published abstracts) that quantitatively measured unmet palliative care needs was also included. Only English-language sources were included. To ensure that the review built on the review by Franks et al. (2000) (12), we only included papers and grey literature evidence published in 2001 or after.

**Table 1.** Inclusion and exclusion criteria for the literature database searches.

|  | Inclusion | Exclusion |
| --- | --- | --- |
| Type of participants | Adults, over 18 with:<br>- Advanced illness<br>- Life-limiting illness<br>- End-stage illness<br>E.g., advanced cancer (stages 3 or 4), end-stage renal disease (stage 5), heart failure (class 3 or 4), advanced dementia (stage 6 or 7)<br>Carers reporting on behalf of adult patients (e.g., bereaved family carers) were included if the remaining inclusion criteria were met. | Adults, over 18 with:<br>- Early stages disease<br>- Survivors<br>- In remission<br>- Not otherwise specified as having advanced illness<br>Children, under 18<br>Family carers only |
| Concept | Unmet palliative/supportive/end-of-life care needs (either as a primary outcome or a secondary outcome), including measures of single care domains of unmet needs, e.g. unmet spiritual care needs.<br>All typologies of unmet needs (felt, expressed, normative, comparative) will be included. | Need for bereavement support only<br>Family carers' unmet support needs only<br>Adult caregivers' perspectives on unmet needs of children or adolescents with advanced illness<br>Focus is solely on psychometric properties of measurements |
| Context | Any geographic location, setting, or health system. | No study was excluded based on context. |
| Type of evidence | Quantitative or mixed-methods original research studies of any design reporting quantitative data<br>Quantitative or mixed-methods systematic or scoping reviews that report quantitative data | Qualitative original research studies<br>Qualitative systematic or scoping reviews<br>Opinion pieces<br>Conference abstracts<br>Published protocols |

### Search strategy

An initial PubMed scoping search of identified relevant published articles and identify target papers. Keywords from titles and abstracts and index terms assigned to the articles from the initial search informed the final search strategy for MEDLINE (Ovid), EMBASE (Ovid), CINAHL (EBSCO) and PsycINFO (Ovid) (see Supplemental file 1). The search strategy was developed in collaboration with an information specialist and adapted for each database and/or information source. Target papers for inclusion in the search strategy were set to check the robustness of the search. All databases were searched on 2 October 2024.

Grey literature was hand-searched by one reviewer (MD) across relevant websites (Hospice UK, National Council for Palliative Care, Marie Curie, Macmillan Cancer Support, The Kings Fund, The Nuffield Trust, RAND and RAND Europe, The Lien Foundation, The Health Foundation, The Strategy Unit and World Health Organisation). Additional grey literature was identified through Open Grey and Google Scholar (using the first 100 references) (21). Forward citation searching of included studies was undertaken using Scopus.

### Study selection

All papers identified from the searches were imported into Covidence and duplicates removed. Titles and abstracts were screened against the inclusion criteria by a single reviewer (TJ/MD/LF/AF), with a subset of 20% papers independently screened by a second reviewer to ensure screening consistency. Full-text screening was conducted by a single reviewer (TJ/MD), with 20% independently screened by a second reviewer (AF/LF) to check agreement. Any disagreements were resolved through discussion with a third reviewer.

### Data extraction and charting

Data from eligible papers and evidence sources were extracted verbatim, where possible, by a single reviewer (TJ/MD/AB) using a bespoke data extraction form in Covidence (see Supplement file 2). A second reviewer (AB/AF) independently checked 20% of the extracted data for accuracy. Any discrepancies between the reviewers were settled through discussion with a third reviewer.

Our extraction form was adapted from the JBI template (21) to fit the review aim, to capture data on study characteristics (e.g. authors, publication year, study design, population); how unmet palliative care needs were defined and measured; and reported (e.g., presentation of study findings of prevalence of unmet needs). For systematic reviews, only review authors’ definitions and operationalisations of unmet palliative care needs, and conclusions regarding its measurement were extracted (excluding primary study results). Data were extracted at study-level; if several papers reported the same study, only one set of data was extracted. Data were recorded and analysed in Microsoft Excel.

## Data analysis

Following scoping review guidance, no quality appraisal was conducted, as the aim was to provide a descriptive overview of the available evidence rather than assess study quality) (23, 24). Data analysis focused on summarising relevant study findings, both by charting and by narrative summary using inductive content analysis (21, 25). Whether studies defined unmet palliative care needs or not was inductively coded as ‘yes’ (a clear explanation of ‘unmet needs’ within a palliative or supportive care context), ‘partially’ (addressing unmet needs, often using prior research findings suggesting issues with palliative care access, but without explicitly delineating the construct) and ‘no’ (no explanation of the construct).

To identify and map the dimensions of unmet needs captured in existing measures, we conducted a framework synthesis (21, 26) based on the domains in Goni-Fuste et al.’s (27) review of comprehensive palliative care needs assessments, encompassing *Physical*, *Psychological*, *Spiritual*, *Social*, *Information*, *Financial*/*legal* (e.g., preparing wills), *Practical* (e.g. household tasks), *Autonomy* (e.g., dependency on others), *Role activities* (e.g., difficulties with employment), *Personal issues* (e.g., handling personal affairs), *Healthcare* (e.g., support from healthcare professionals). Domains were added as needed (26).

## Results

The searches and study inclusion are presented in a PRISMA flow diagram in Figure 1 (28). The database searches identified a total of 4,513 sources after duplicates were removed. Following the title and abstract screening, 274 sources were sought for full-text review. After full-text screening against inclusion and exclusion criteria, 70 studies (reported in 74 peer-reviewed scientific papers, and in one grey literature source) were included.

**Figure 1.**
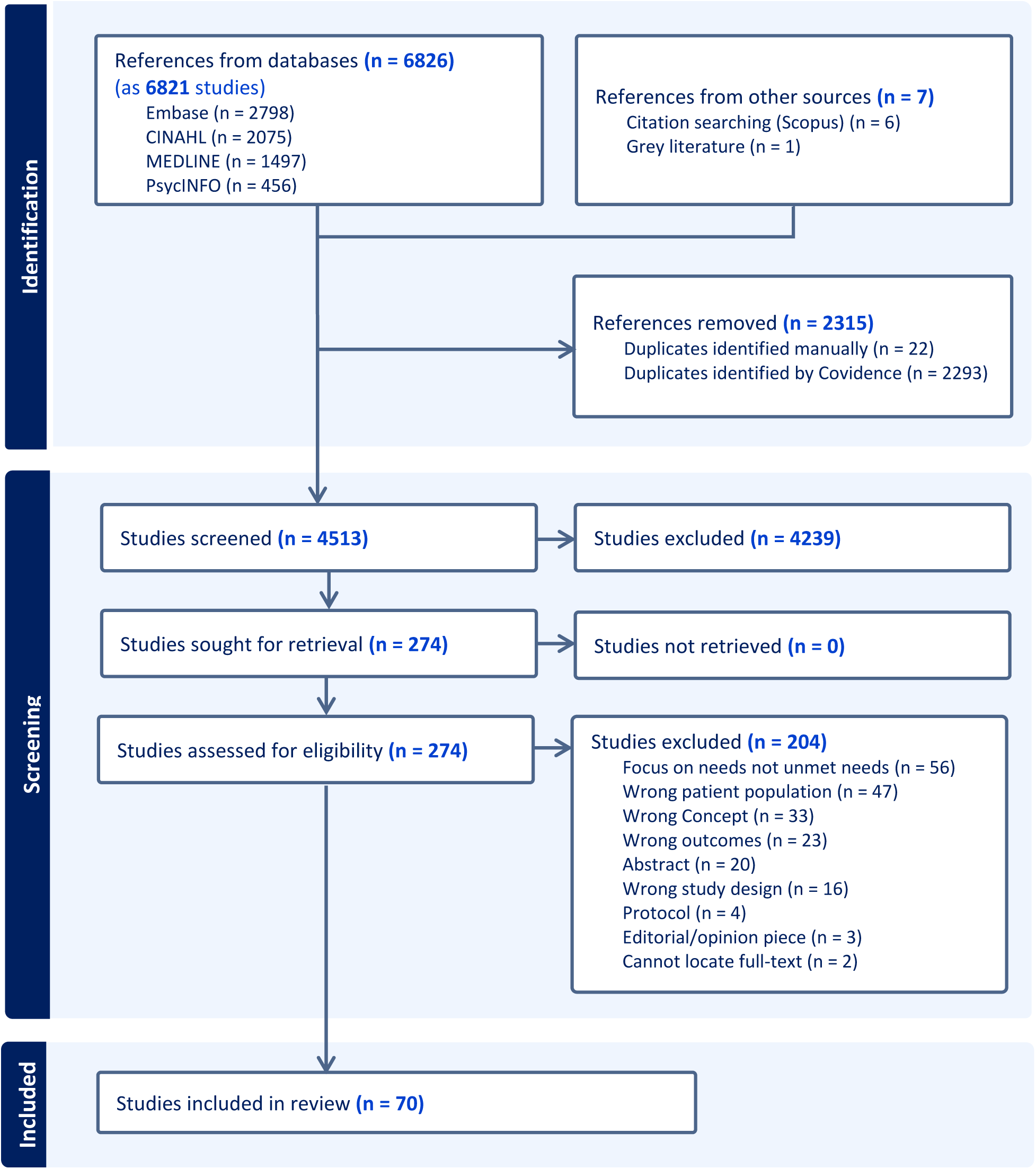
PRISMA flow diagram of study selection.

The included studies were published between 2001-2024. In total, 61 were primary evidence studies (i.e., original research) (29–90), and nine were reviews (91–99). The original studies (n=61) were conducted in 16 countries, primarily Western countries such as the United States (n=23), Australia (n=9), Canada (n=5), and the United Kingdom (n=5).

### Study descriptives

#### Study designs

Details of study characteristics are presented in Table 2. Most included studies used observational designs: commonly cross-sectional (n=43, 59%), prospective cohort (n=8, 11%), and retrospective cohort (n=6, 9%) designs. The primary evidence studies that directly measured unmet palliative care needs, mostly used convenience samples (n=49, 80%). Only two primary evidence studies reported on patient and public involvement (PPI): one mentioned PPI input when developing the measure (86), and one other pilot-tested their questionnaire to ensure it was not too distressing (31).

**Table 2.** Characteristics of included studies (n=70), ordered by study design.

| Authors and year | Country | Summary of study aim | Study design | Population | Study setting | Sample | Recruitment | Sample size |
| --- | --- | --- | --- | --- | --- | --- | --- | --- |
| **Prospective cohort studies** |  |  |  |  |  |  |  |  |
| Faris et al.<sup>1</sup> (2024) | Australia | Assess unmet needs among patients with brain tumours and explore feasibility of addressing these in clinical practice | Prospective cohort study | Advanced cancer | Hospital | Convenience sample | Clinic patients | 116 |
| Hamano et al. (2023) | Japan | Assess unmet needs among non-cancer patients across the illness trajectory and factors related to these | Prospective cohort study (multi-centre) | Unspecified life-limiting illness | Care home | Convenience sample | Clinic patients | 785 (T0); 553 (T1); 370 (T2); 317 (T3) |
| Hampton et al. (2007) | United States | Assess spiritual needs among advanced cancer patients and whether they were met | Prospective cohort study | Advanced cancer | Hospice | Convenience sample | Clinic patients | 90 |
| Kiely et al. (2010) | United States | Assess how bereaved family member's experiences of hospice care is associated with symptom treatment and unmet care needs in the last days of life | Prospective cohort study | Dementia | Care home | Convenience sample | Clinic patients | 323 |
| Kirkland et al.<sup>2</sup> (2021) | Canada | Compare characteristics between patients who have unmet palliative care needs and those who do not | Prospective cohort study | Unspecified life-limiting illness | Hospital | Convenience sample | Clinic patients | 663 |
| Makaroun et al. (2018) | United States | Examine how late healthcare transitions are associated with unmet needs, communication and quality of care | Prospective cohort study | Decedents (any site of death and cause of death) | Any care setting | Nationally representative sample | National Health and Aging Trends Study register | 1,653 |
| Sharpe et al. (2005) | Australia | Assess unmet needs in a patient group and explore if ratings differ between patients, their families, and healthcare professionals | Prospective cohort study | Advanced cancer | Hospital | Convenience sample | Clinic patients | 142 |
| Vogt et al. (2021) | Germany | Assess symptom burden and palliative care needs during the first year of diagnosis with incurable cancer | Prospective cohort study | Advanced cancer | Hospital (cancer centre) | Convenience sample | Clinic patients | 161 |
| **Retrospective cohort studies** |  |  |  |  |  |  |  |  |
| Arabadzhyan et al. (2024) | United Kingdom | Explore unmet care need in the final months of life among people who died of cancer during the Covid-19 pandemic | Retrospective cohort study | Advanced cancer | Any care setting | Population-based sample | National mortality register | 130,490 |
| Axelsson et al. (2018) | Sweden | Assess symptom prevalence, relief, and management among patients with end-stage kidney disease | Retrospective cohort study | End-stage renal disease | Any care setting | Population-based sample | National palliative register | 472 |
| Hildenbrand et al. (2022) | United States | Explore the association between cancer patients' distress and urgent healthcare use | Retrospective cohort study | Advanced cancer | Hospital (cancer centre) | Convenience sample | Clinic patients | 848 |
| Hwang et al. (2019) | United States | Characterise the outcomes for patients with end-stage renal disease, define patterns of palliative care use and identify unmet needs | Retrospective cohort study | End-stage renal disease | Hospital | Convenience sample | Medical records from patient register | 9,363 |
| Safabakhsh et al. (2023) | United States | Calculate unmet need for palliative care among hospitalised patients | Retrospective cohort study | Unspecified life-limiting illness | Hospital | Population-based sample | Clinic patients | 92,047 |
| Westley-Wise et al. (2022) | Australia | Assess the unmet need for palliative and end-of-life care, and explore sociodemographic and diagnostic factors associated with suboptimal access to these services | Retrospective cohort study | Decedents (any site of death and cause of death) | Hospital: Community health centre | Population-based sample | Medical records from patient register | 3,175 |
| **Experimental study** |  |  |  |  |  |  |  |  |
| Paterson et al. (2018) | United Kingdom | Explore the effect of an intervention on unmet needs among advanced prostate cancer patients | Randomised controlled trial | Advanced cancer | Hospital | Random sample | Clinic patients | 47 |
| **Mixed-methods studies** |  |  |  |  |  |  |  |  |
| Paterson et al.(2017) | United Kingdom | Assess unmet care needs among advanced prostate cancer patients | Mixed-methods study | Advanced cancer | Hospital (cancer centre) | Convenience sample | Clinic patients | 31 |
| Spence et al. (2020) | Canada | Describe the symptom burden of patients during consultation with an integrated palliative radiotherapy service | Mixed-methods study | Advanced cancer | Hospital (cancer centre) | Convenience sample | Clinic patients | 607 |
| Teno et al. (2021) | United States | Examine bereaved relatives' perceptions of care received in the last month of life | Mixed-methods study | Deaths from any non-traumatic causes | Any care setting | Convenience sample | Mail | 623 |
| **Cross-sectional studies** |  |  |  |  |  |  |  |  |
| Abu-Odah et al. (2022) | Palestine | Explore cancer patients' needs | Cross sectional study | Advanced cancer | Hospital | Convenience sample | Clinic patients | 379 |
| Alnajar et al. (2024) | Jordan | Assess and compare unmet needs between cancer and non-cancer patients | Cross sectional (comparative) study | Advanced cancer; Non-cancer chronic conditions | Hospital | Convenience sample | Clinic patients | 458 |
| Anderson et al. (2001) | United Kingdom | Determine the main concerns experienced by patients with terminal illness and if they were met | Cross sectional study | Palliative care patients (unspecified life-limiting illness); Heart failure | Palliative care services (hospital, hospice, & community-based); Hospital (outpatient clinic) | Convenience sample | Clinic patients | 279 |
| Aranda et al. (2005) | Australia | Identify patterns of support and information needs among cancer patients | Cross sectional study | Advanced cancer | Hospital | Convenience sample | Clinic patients | 105 |
| Blindbaek et al. (2014) | Denmark | Measure prevalence of problems experienced by patients with advanced kidney disease and whether these were met | Cross sectional study | End-stage renal disease | Hospital | Convenience sample | Clinic patients | 27 |
| Buzgova et al. (2014) | Czech Republic | Determine associations between unmet needs, quality of life, anxiety and depression among cancer patients | Cross sectional study | Advanced cancer | Hospital | Convenience sample | Clinic patients | 93 |
| Buzgova et al.<sup>3</sup> (2016) | Czech Republic | Identify unmet palliative care needs among hospitalised patients and explore factors related to these | Cross sectional study | Unspecified life-limiting illness | Hospital | Convenience sample | Clinic patients | 349 |
| Chuang et al. (2017) | United States | Estimate generalist and specialist palliative care delivery, estimate unmet needs among inpatients and explore ethnic disparities in palliative care delivery | Cross sectional study | Unspecified life-limiting illness | Hospital | Convenience sample | Clinic patients | 8,301 (sub-sample (n=401) for in-depth analysis) |
| Connor et al. (2005) | United States | Assess differences in care among hospice providers | Cross sectional study | Hospice decedents (any cause of death) | Hospice | Convenience sample | Mail | 29,292 |
| Cooper et al. (2021) | Australia | Determine how many hospital inpatients would benefit from palliative care and how many were receiving it | Cross sectional (prevalence) study | Unspecified life-limiting illness | Hospital | Convenience sample | Clinic patients | 270 |
| Cooper et al. (2024) | Australia | Determine how many hospital inpatients would benefit from palliative care and how many were receiving it | Cross sectional (prevalence) study | Unspecified life-limiting illness | Hospital | Convenience sample | Clinic patients | 279 |
| Currow et al. (2008) | Australia | Determine and compare perceived unmet palliative care needs across four subpopulations: those with non-cancer diagnoses; those 75 years and older; those with lower household incomes; and those of non-English speaking backgrounds | Cross sectional (comparative) study | Unspecified life-limiting illness | Specialist palliative care service | Nationally representative sample | Post-bereavement study | 7,105 |
| DeGroot et al. (2023) | United States | Examine palliative care needs among patients with heart failure | Cross sectional study | Heart failure | Hospital | Convenience sample | Clinic patients | 286 |
| Driessen et al. (2023) | the Netherlands | Gain insight into disease- and care-related needs among patients with advanced cancer and explore factors related to these | Cross sectional study | Advanced cancer | Hospital | Convenience sample | Clinic patients | 1,103 |
| Effendy et al. (2015) | Indonesia and the Netherlands | Determine and compare unmet needs between patients with advanced cancer in Indonesia and the Netherlands | Cross sectional study | Advanced cancer | Hospital; Care home | Convenience sample | Clinic patients | 274 |
| Hasegawa et al. (2016) | Japan | Assess unmet needs among cancer patients and factors related to these | Cross sectional study | Advanced cancer | Hospital | Convenience sample | Clinic patients | 45 |
| Hasegawa et al. (2021) | Japan | Assess unmet rehabilitation needs among hospice inpatients and how this related to medico-psychosocial factors | Cross sectional study | Advanced cancer | Palliative care service | Random sample | Post-bereavement study | 416 |
| Huang et al. (2020) | Singapore | Assess care needs, functional status and quality of life among lung cancer patients aged 50 and older | Cross sectional study | Advanced cancer | Hospital (cancer centre) | Convenience sample | Clinic patients | 103 |
| Husain et al. (2013) | Canada | Describe continuity of care and care needs among advanced lung cancer patients and factors related to these | Cross sectional study | Advanced cancer | Hospital | Convenience sample | Clinic patients | 116 |
| Hwang et al. (2004) | United States | Assess unmet needs among advanced cancer patients and factors related to these | Cross sectional study | Advanced cancer | Hospital | Convenience sample | Clinic patients | 296 |
| Jeyasingam et al. (2008) | Australia | Assess unmet ADL needs among inpatients in a | Cross sectional | Palliative care inpatients | Palliative care service | Convenience sample | Clinic patients | 30 patient- |
|  |  | palliative care unit and explore congruence ratings by patients, families and care professionals | (prevalence) study | (any life-limiting illness) |  |  |  | caregiver dyads |
| Johnsen et al. (2013) | Denmark | Assess the adequacy of help from healthcare services to resolve problems experienced by patients with advanced cancer | Cross sectional study | Advanced cancer | Hospital | Nationally representative sample | Medical records from patient register | 977 |
| Kavalieratos et al. (2014) | United States | Assess and compare unmet needs between community-dwelling patients with heart failure and cancer | Cross sectional (comparative) study | Advanced cancer; Heart failure | Palliative care service | Convenience sample | Clinic patients | 1,031 |
| Khan et al. (2012) | Canada | Assess unmet needs among palliative radiotherapy patients and explore if ratings differ between patients and their families | Cross sectional study | Advanced cancer | Hospital (cancer centre) | Convenience sample | Clinic patients | 40 |
| Miniotti et al. (2019) | Italy | Assess care needs, quality of life, and psychological morbidity of patients with advanced colorectal cancer | Cross sectional study | Advanced cancer | Hospital | Convenience sample | Clinic patients | 203 |
| Munn et al. (2006) | United States | Explore if hospice care for nursing home residents is associated with symptom management, personal care, spiritual support, and care satisfaction. | Cross sectional study | Care home residents (unspecified life-limiting illness) | Care home | Convenience sample | Phone | 124 |
| Oh et al. (2019) | South Korea | Assess unmet needs among patients with amyotrophic lateral sclerosis (ALS) and determine its relation to functional status and quality of life | Cross sectional study | ALS | Hospital | Convenience sample | Clinic patients | 186 |
| Osse et al. (2005) | the Netherlands | Assess needs among cancer patients and whether these were met | Cross sectional study | Advanced cancer | Any care setting | Convenience sample | Clinic patients | 94 |
| Park et al. (2017) | United States | Assess spiritual needs among heart failure patients | Cross sectional study | Heart failure | Hospital (outpatient clinic) | Convenience sample | Clinic patients | 111 |
| Pearce et al. (2012) | United States | Assess the prevalence of insufficient spiritual care and its association with patient-reported outcomes | Cross sectional study | Advanced cancer | Hospital | Convenience sample | Clinic patients | 150 |
| Rachakonda et al. (2015) | Australia | Assess unmet needs in cancer patients living in rural areas. | Cross sectional study | Advanced cancer | Community (rural care centre) | Convenience sample | Clinic patients | 75 |
| Rhodes et al. (2012) | United States | Examine bereaved family members' satisfaction with hospice care depending on the proportion of African American patients in hospice. | Cross sectional study | Hospice decedents (any cause of death) | Palliative care service (hospice) | Convenience sample | Mail | 11,892 |
| Schenker et al. (2014) | United States | Examine the association between unmet needs and interest in seeing a palliative care physician | Cross sectional study | Advanced cancer | Palliative care service (cancer centre with palliative care clinic) | Convenience sample | Clinic patients | 169 |
| Strupp et al. (2018) | Germany | Characterise symptoms of people severely affected by Parkinson Disease/Atypical Parkinsonism and describe their prevalence of unmet needs | Cross sectional study | Parkinson's Disease | Any care setting | Convenience sample | Magazine advertisement | 814 |
| Szekendi et al. (2016) | United States | Estimate the need for a palliative care referral among hospital inpatients, determine the proportion of who receives a referral for palliative care consultation, explore organizational characteristics influencing referral | Cross sectional (prevalence) study | Unspecified life-limiting illness | Hospital | Random sample | Clinic patients | 2,119 |
| Teno et al. (2004) | United States | Evaluate the dying experience at home and in institutional settings | Cross sectional study | People who died from any chronic illness | Any care setting | Nationally representative sample | Post-bereavement study | 1,578 |
| Teno et al. (2007) | United States | Examine the association of perceived quality of care in hospice with length of stay and timing of referral | Cross sectional study | Hospice decedents (any cause of death) | Palliative care service (hospice) | Convenience sample | Mail | 106,514 |
| Teno et al. (2011) | United States | Examine the perceived effectiveness of hospice services for people dying with dementia | Cross sectional study | Dementia | Any care setting | Random sample | Phone | 538 |
| Trandel et al. (2019) | United States | Determine the prevalence of existential distress by identifying unmet existential needs and explore associations with symptom burden | Cross sectional study | Cystic fibrosis | Hospital (outpatient clinic) | Convenience sample | Clinic patients | 164 |
| Wang et al. (2021) | China | Examine palliative care needs among advanced cancer patients and their informal caregivers | Cross sectional study | Advanced cancer | Hospital | Convenience sample | Clinic patients | 428 |
| Wang et al. (2023) | China | Identify palliative care needs of patients with end-stage renal disease undergoing maintenance haemodialysis, and explore the influencing factors of unmet needs | Cross sectional study | End-stage renal disease | Hospital (outpatient clinic) | Convenience sample | Clinic patients | 305 |
| Watson et al. (2019) | United Kingdom | Examine experiences of care and care needs among pancreatic cancer patients | Cross sectional study | Advanced cancer | Community (NHS centre) | Convenience sample | Clinic patients | 274 |
| Wegier et al. (2021) | Canada | Measure the prevalence of unmet palliative needs among patients | Cross sectional study | People estimated to be in their final | Hospital (quaternary) | Convenience sample | Clinic patients | 403 |
|  |  | estimated to be in their last year of life |  | year of life (any life-limiting illness) | healthcare facility) |  |  |  |
| **Systematic reviews** |  |  |  |  |  |  |  |  |
| Bore et al. (2024) | Review conducted in Australia | Determine the prevalence of unmet care needs among cancer patients and explore factors contributing to these | Systematic review | Advanced cancer | Any care setting in Sub-Saharan Africa | - | - | - |
| Chen et al. (2020) | United States | Identify ways to measure unmet needs for symptom management and communication in a palliative population | Systematic review | Unspecified life-limiting illness | Not stated | - | - | - |
| Fu et al. (2020) | Review conducted in Australia | Assess care and unmet care needs and factors related to these | Systematic review | Advanced cancer | Not stated | - | - | - |
| Harrison et al. (2009) | Review conducted in Australia | Assess unmet needs among cancer patients across the illness trajectory, explore associated factors and describe study designs used to research this | Systematic review | Advanced cancer | Any care setting | - | - | - |
| Hart et al. (2022) | Review conducted in Australia | Assess unmet needs among advanced cancer patients | Systematic review | Advanced cancer | Not stated | - | - | - |
| Moghadam et al. (2016) | Review conducted in the United Kingdom | Assess unmet needs among advanced cancer patients, identifying domains of care and instruments to measure unmet need. | Systematic review | Advanced cancer | Not stated | - | - | - |
| Schmidt et al. (2023) | Review conducted in Switzerland | Identify validated tools for clinicians to capture patient reports on unmet needs, to guide palliative care interventions | Systematic review | Unspecified life-limiting illness | Any care setting | - | - | - |
| Ventura et al. (2014) | Review conducted in Australia | Summarise the literature on unmet needs of palliative home care patients and carers | Systematic review | Unspecified life-limiting illness | Palliative care service (home care) | - | - | - |
| Wang et al. (2018) | Review conducted in Hong Kong | Identify the unmet care needs and their associated factors in patients with advanced cancer and their informal caregivers, and summarise the tools used in the included studies | Systematic review | Advanced cancer | Any care setting | - | - | - |

Notes: <sup>1</sup> Study also reported in Halkett et al. 2015; <sup>2</sup> Study also reported in Kirkland et al. 2022 & Kruhlak et al., 2021; <sup>3</sup> Study also reported in Buzgova et al., 2015

### Populations and settings

As shown in Table 2, most studies (n=45, 64%) focused on unmet needs within a single disease group, commonly advanced cancer (n=36, 51%), with three (3%) studies comparing two groups. One-third of studies (n=23, 33%) included patients or decedents with any life-limiting illness. Study settings varied: most studies used hospital settings (n=40,57%) (including cancer centres and outpatient clinics), whereas 11 (16%) focused on palliative care services (including hospices, hospital units, and home care). Eleven (16%) studies included participants from any care setting, and four (6%) did not specify a setting.

### Definitions of unmet palliative care needs

Details of how studies defined, measured, and reported unmet palliative care needs are presented in Table 3. Only 11 (16%) studies explicitly provided a definition of unmet palliative care needs, whereas 23 (33%) partially defined the concept – often with reference to prior research on care needs. More than half (n=36, 51%) provided no definition.

**Table 3.**
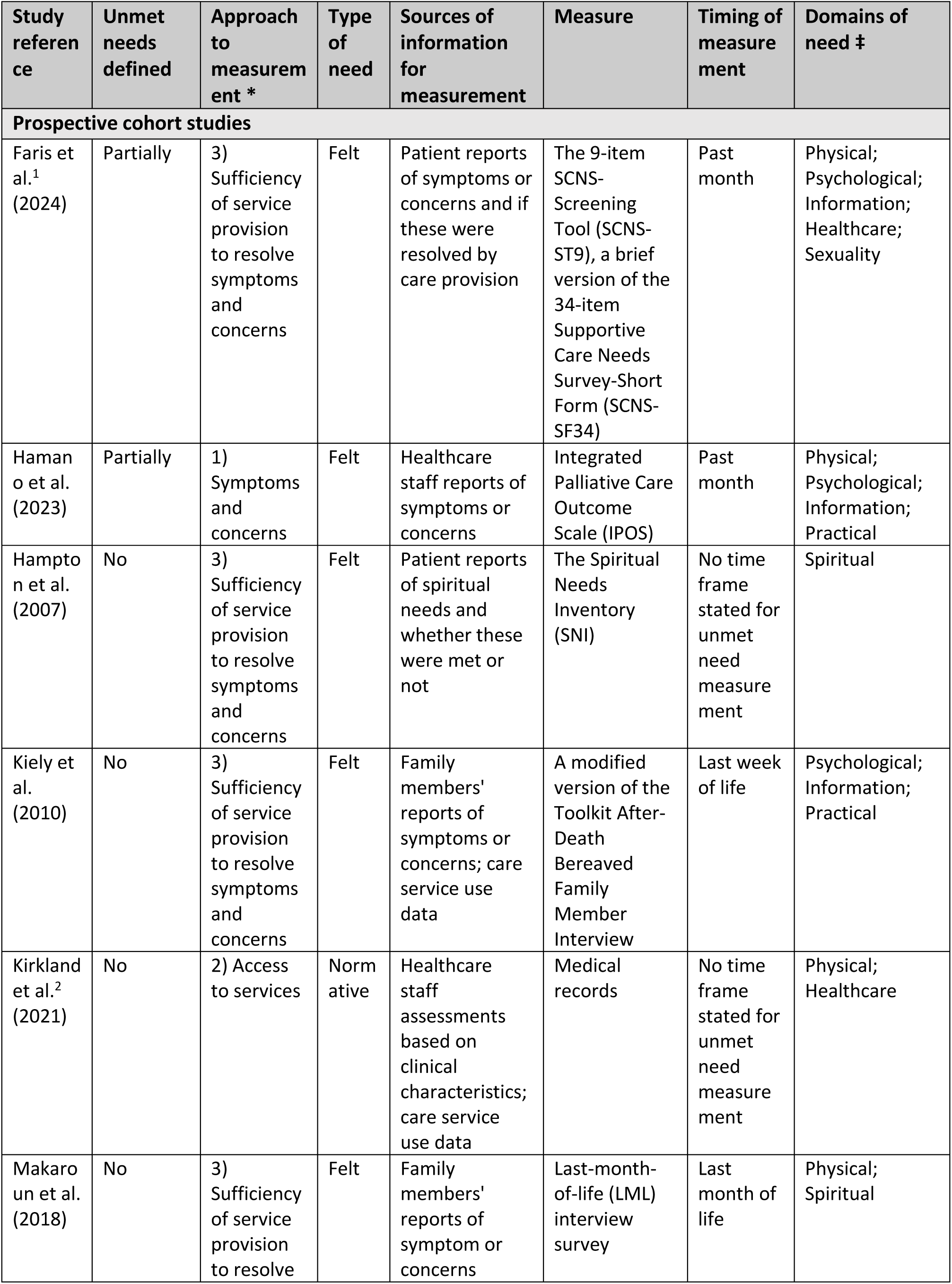
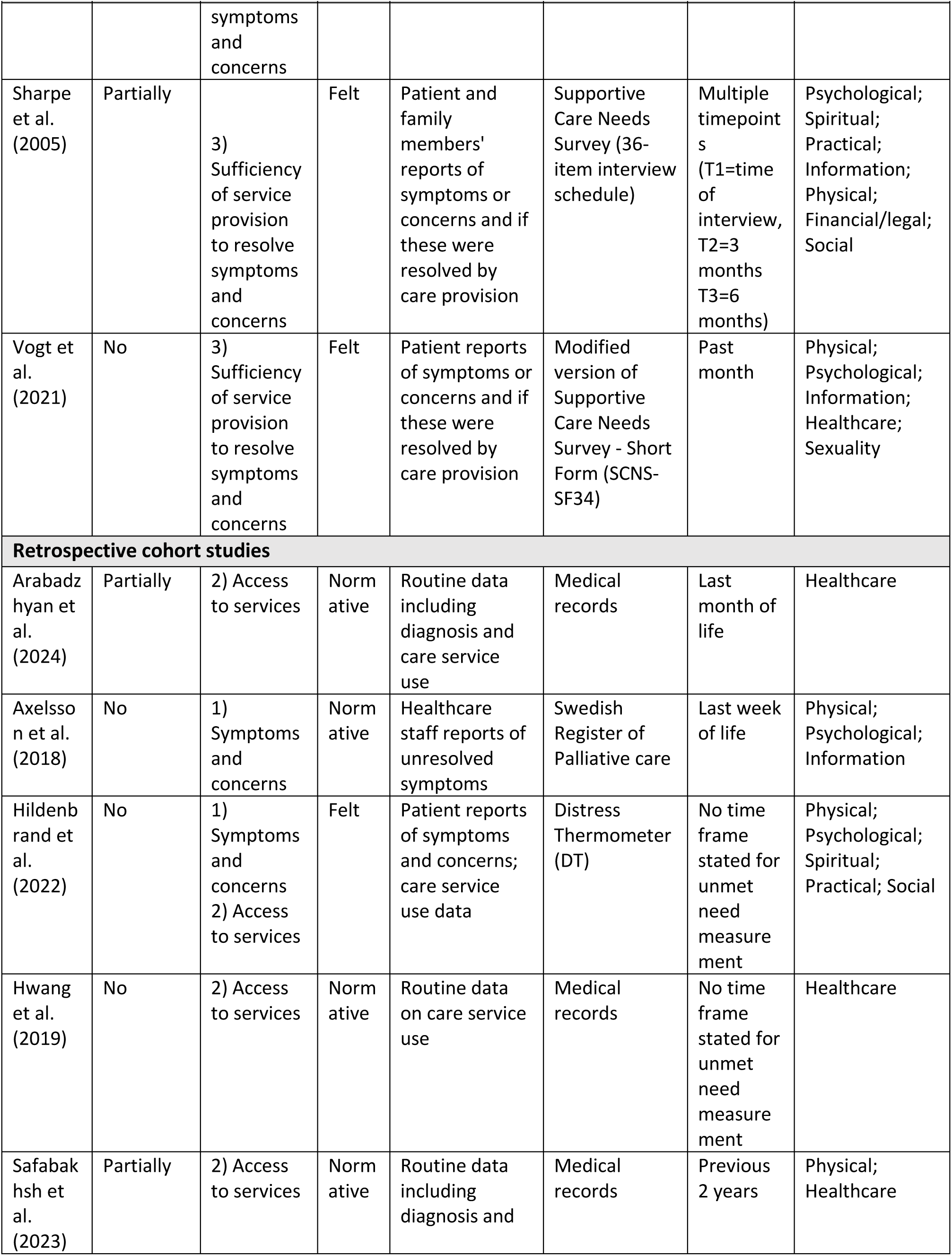
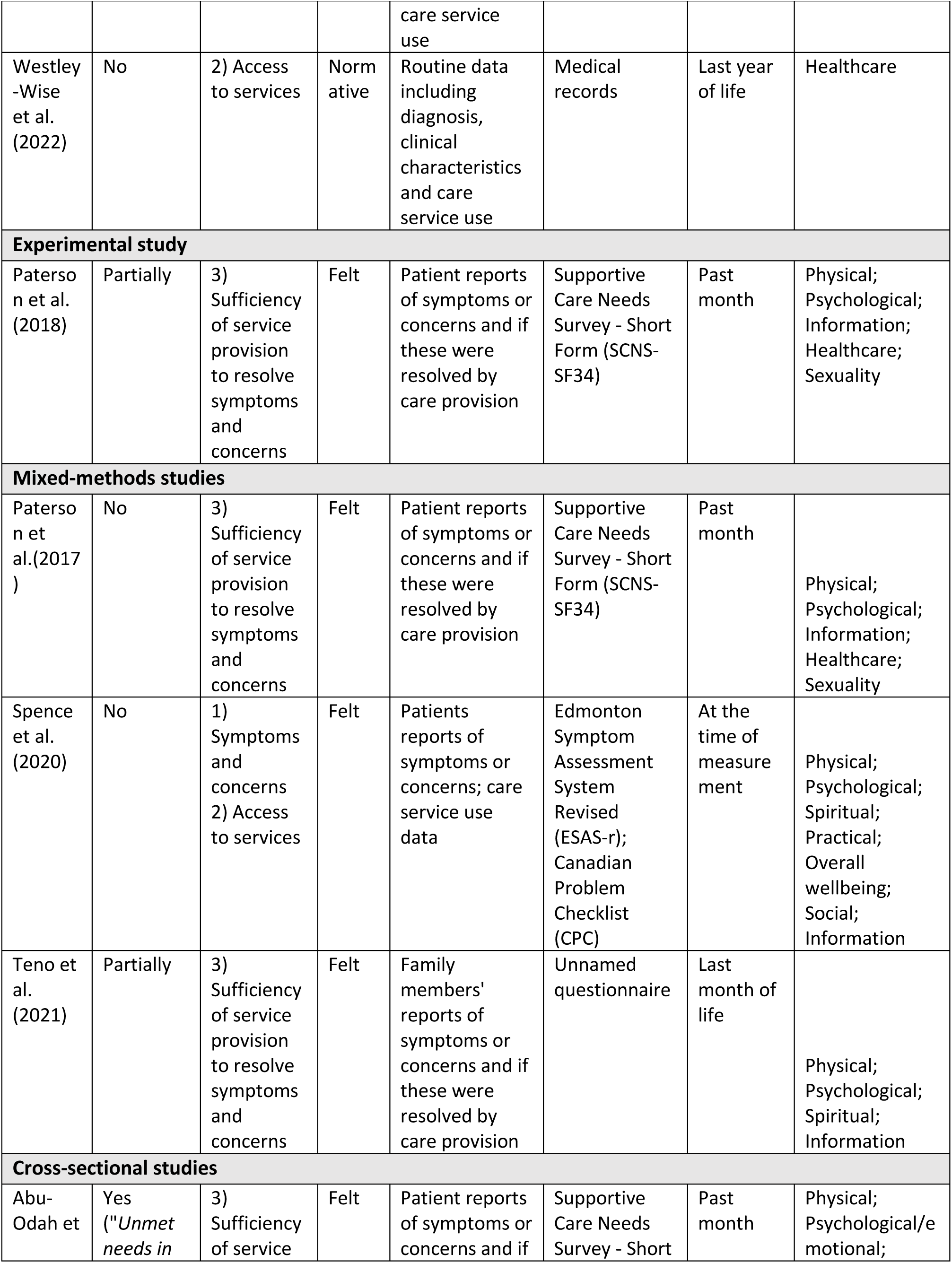
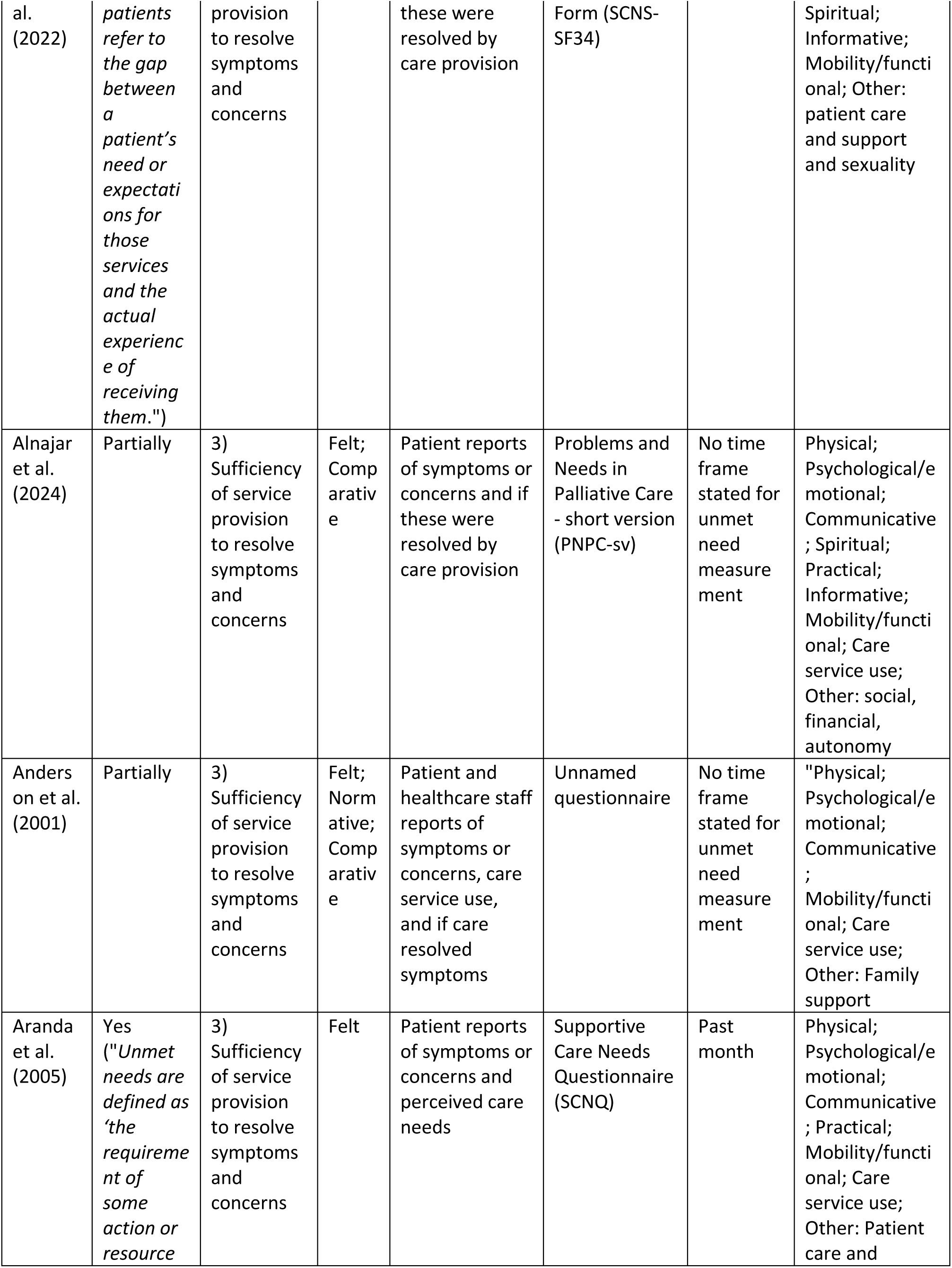
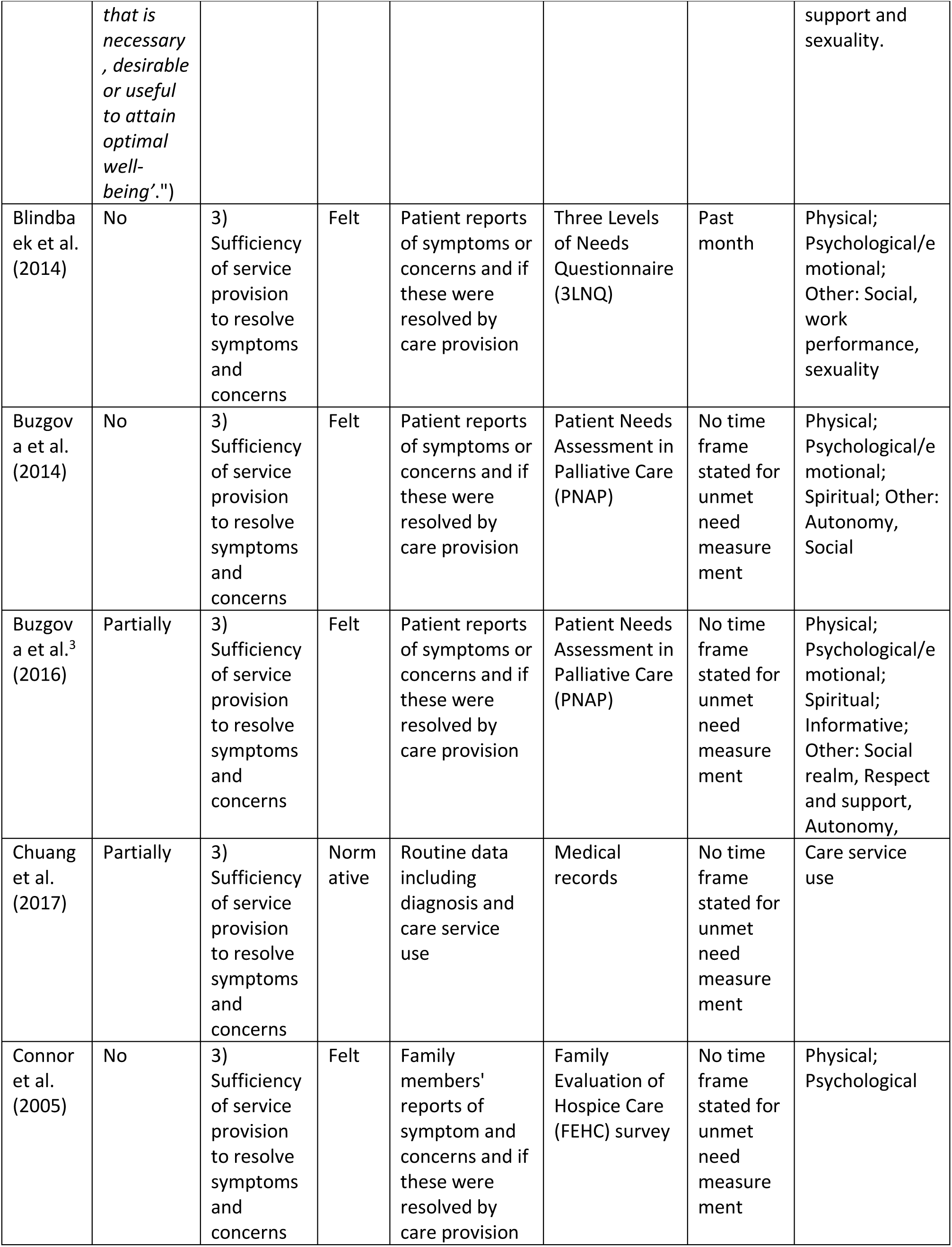
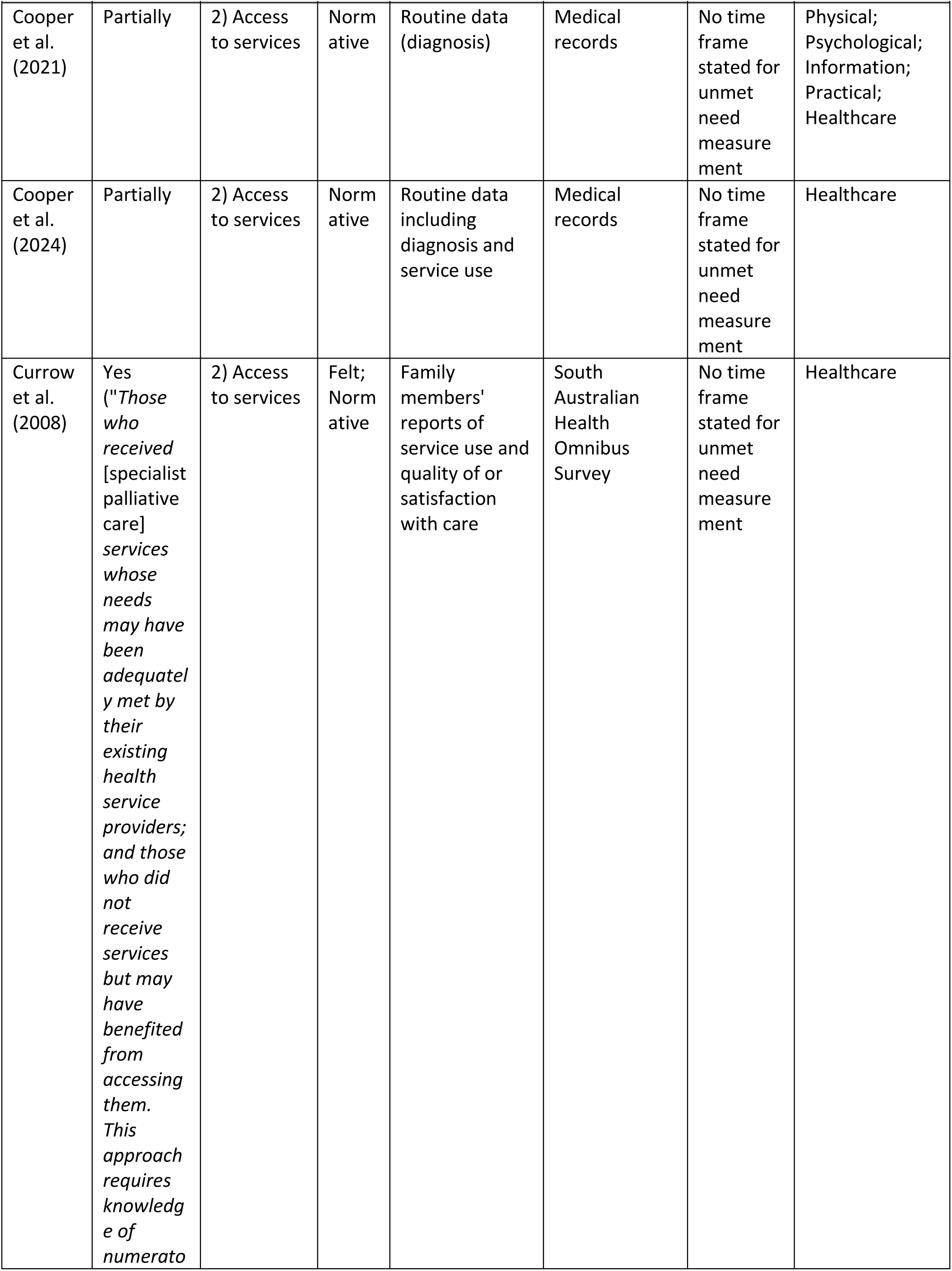
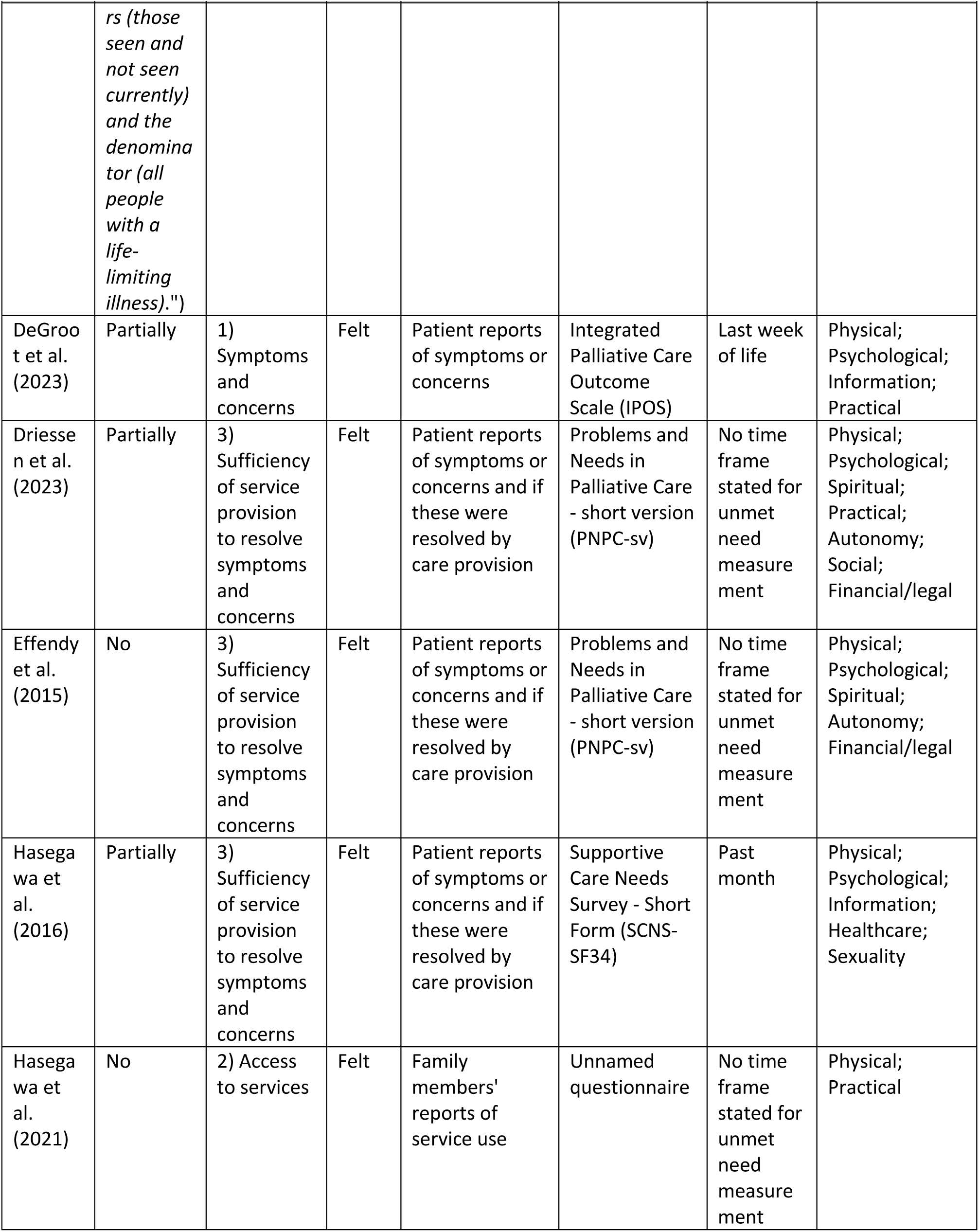
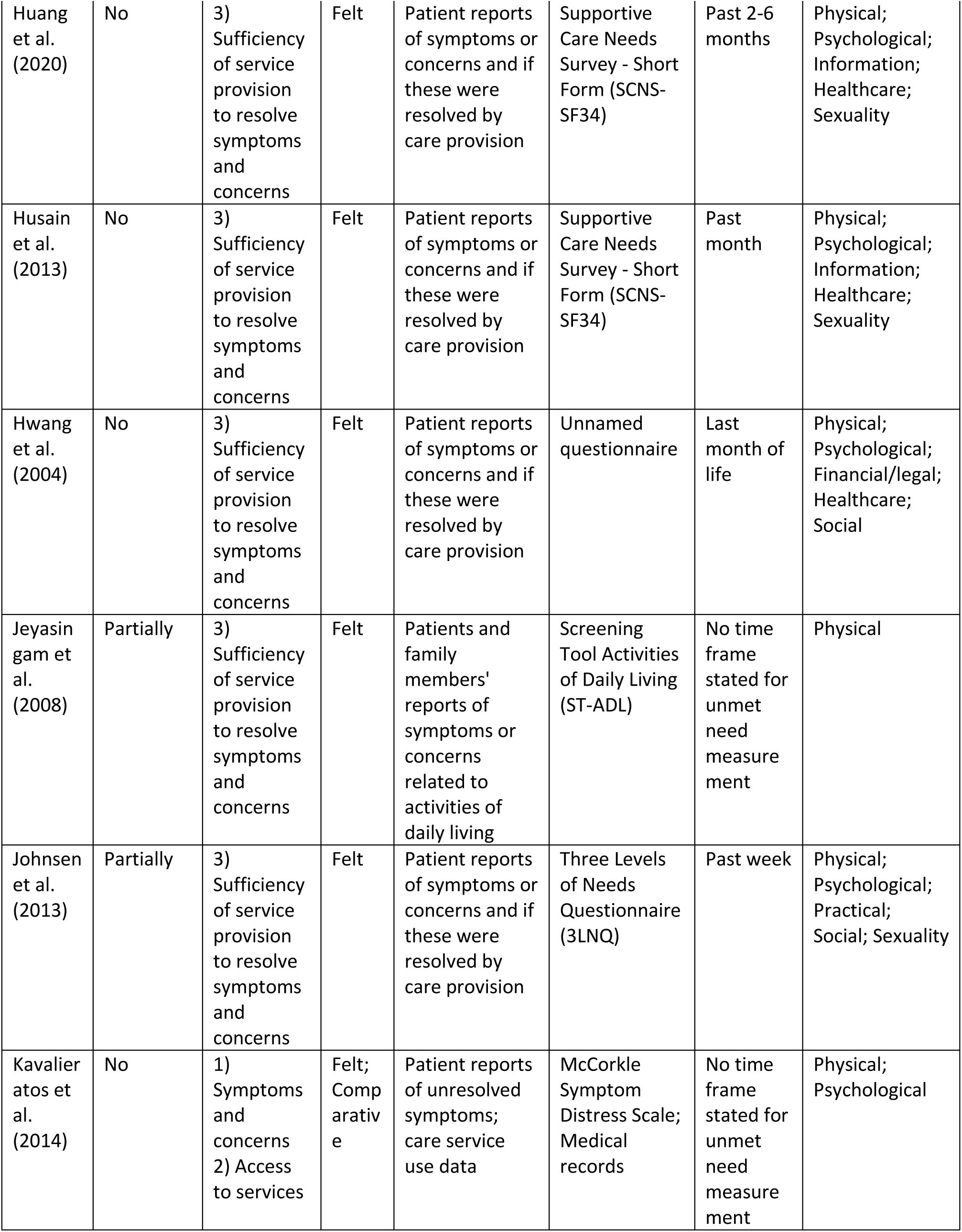
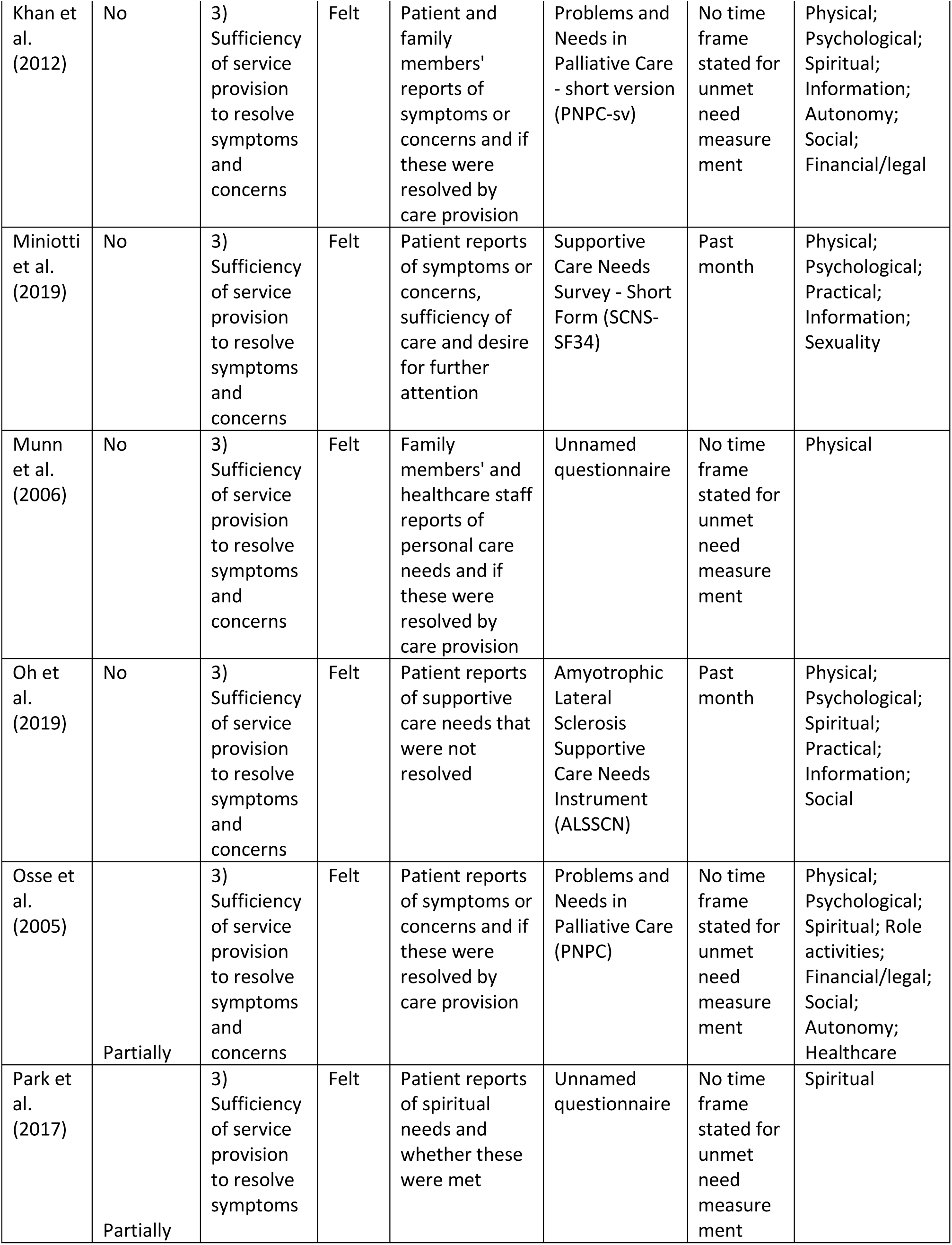
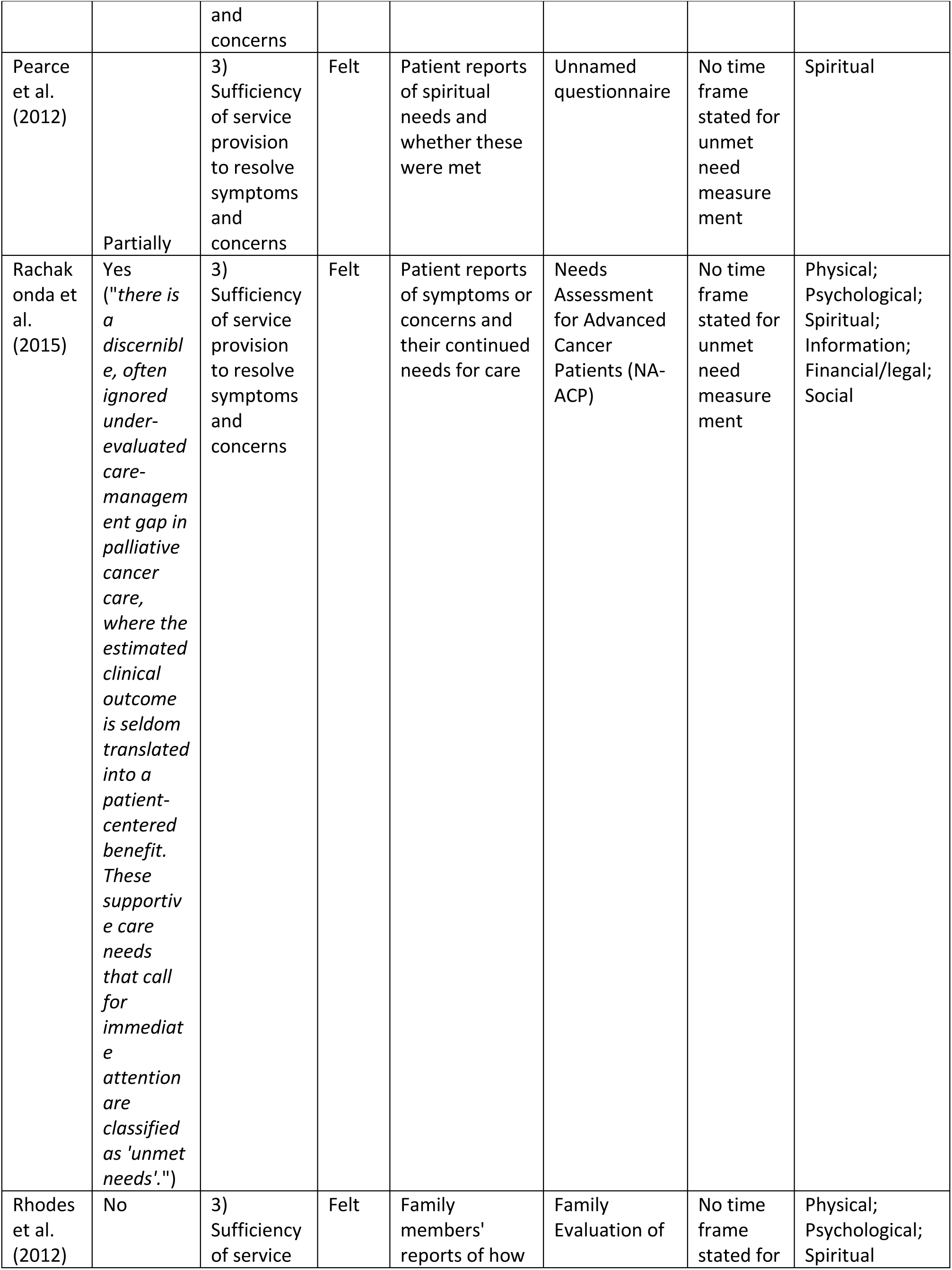
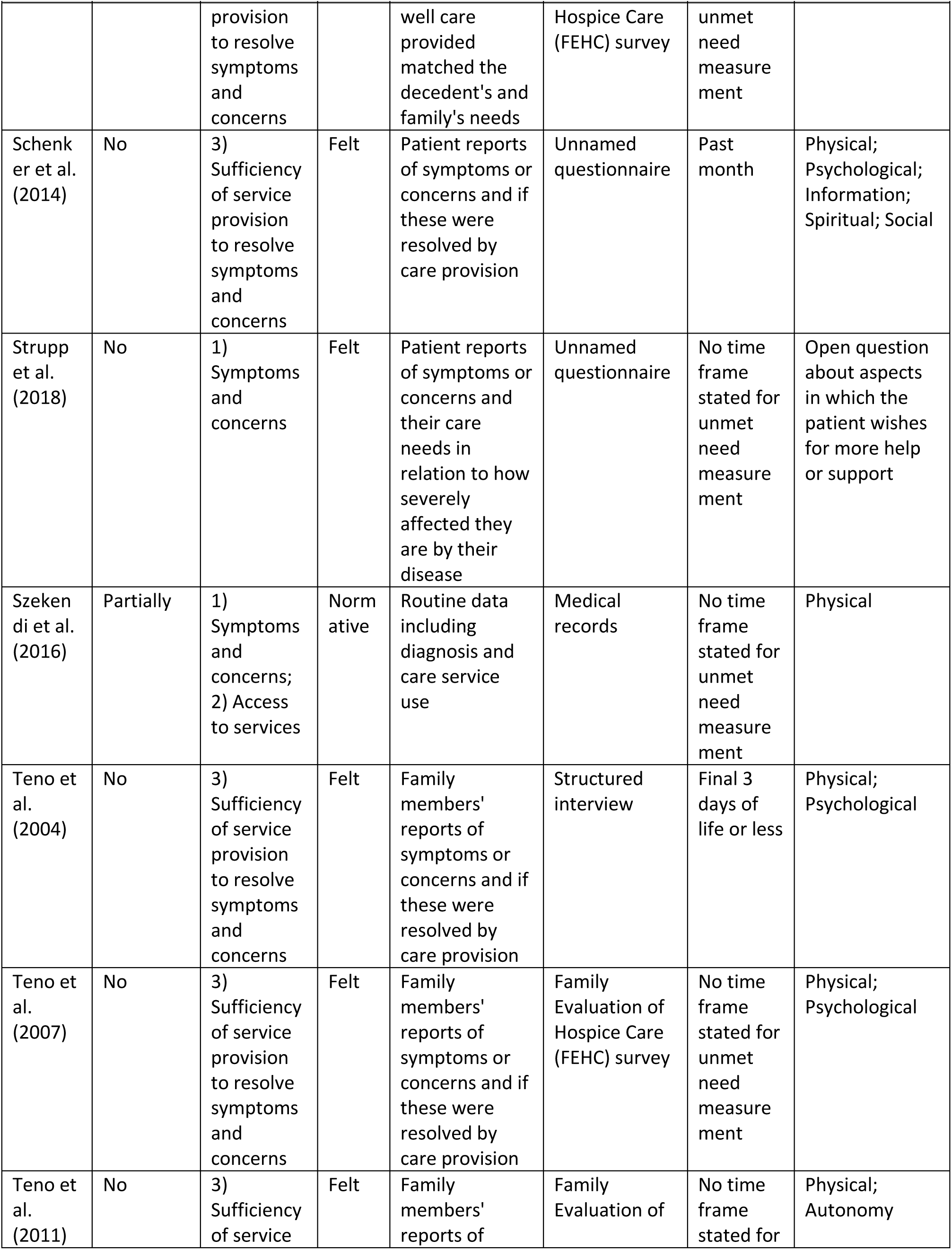
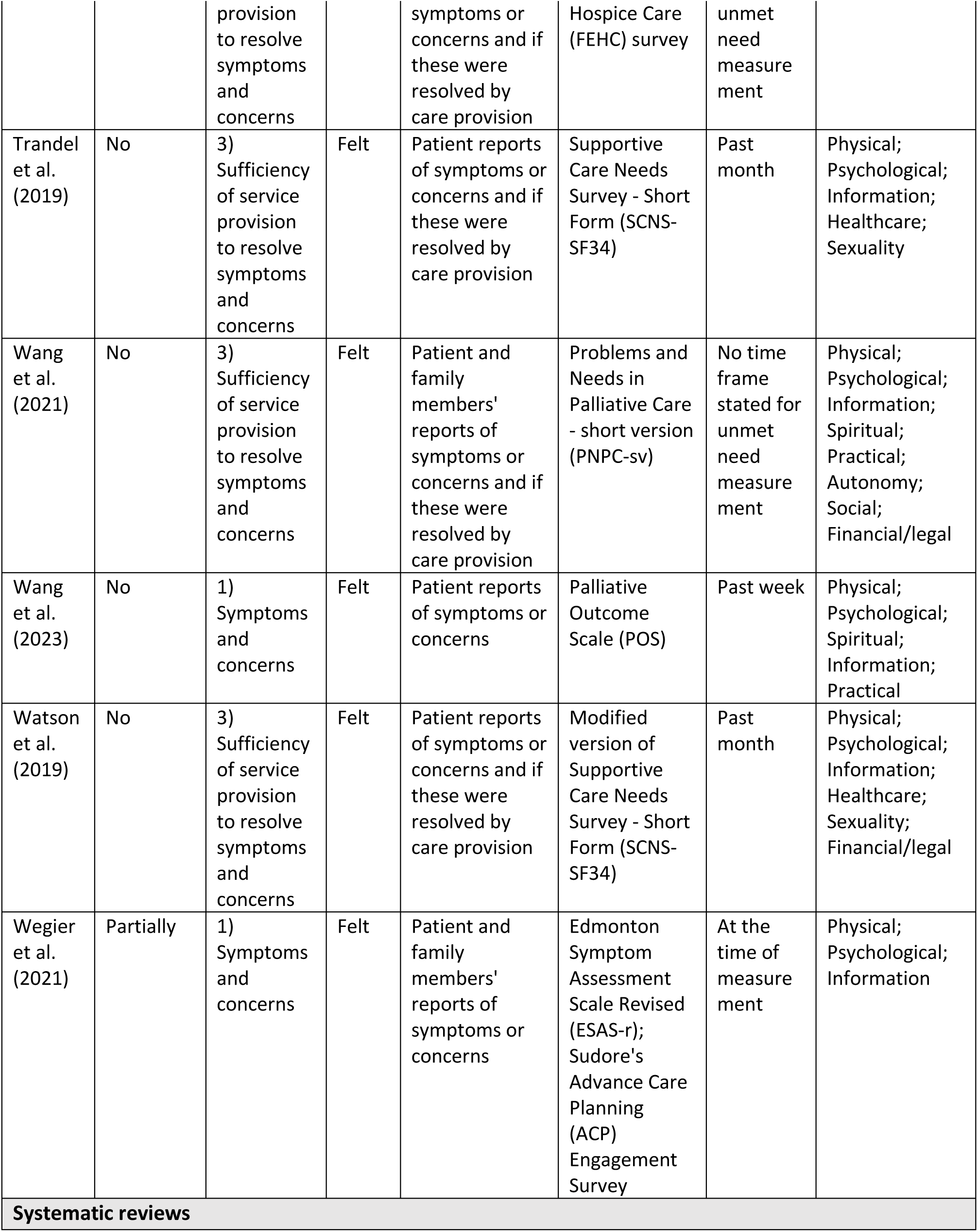
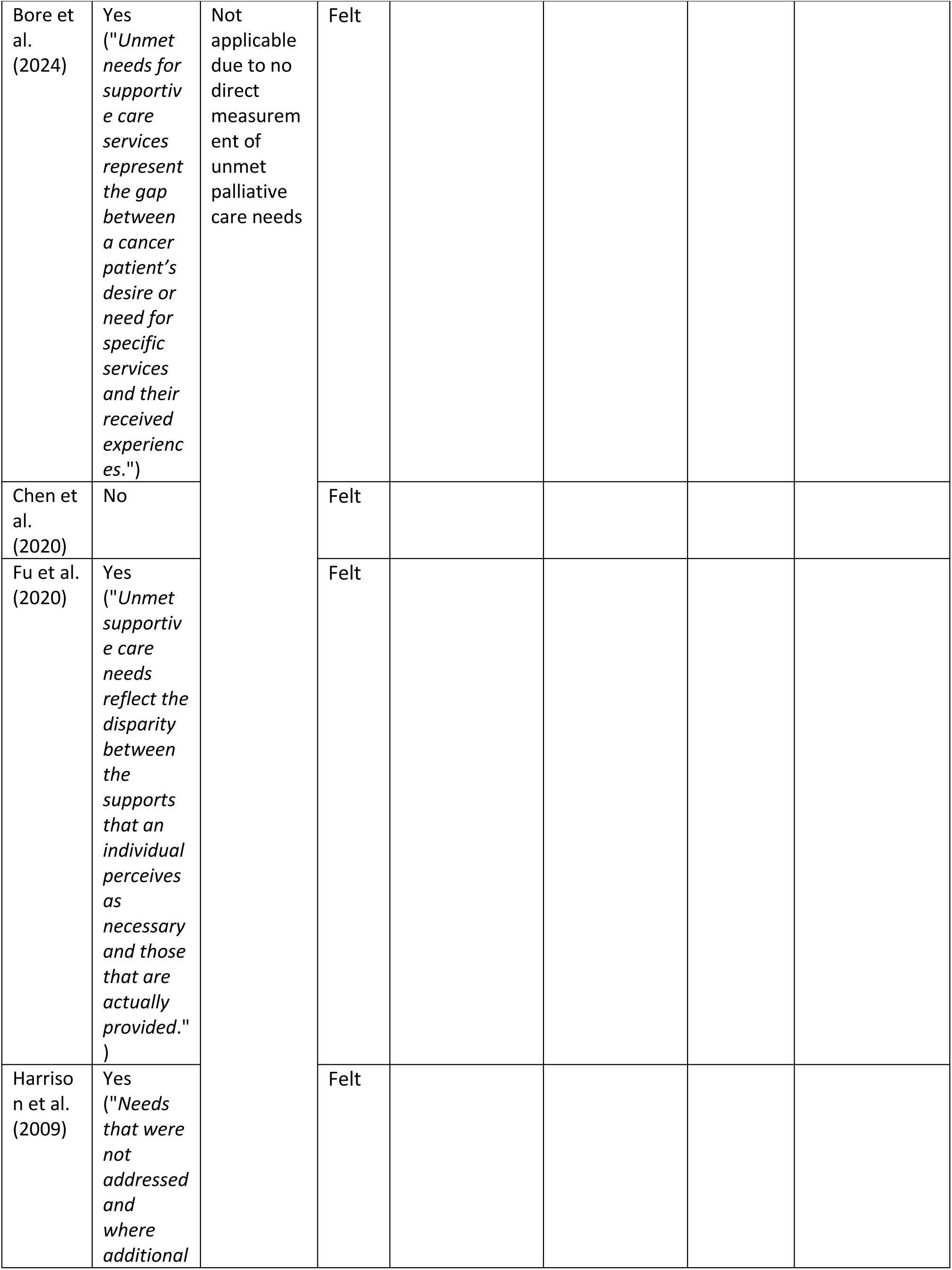
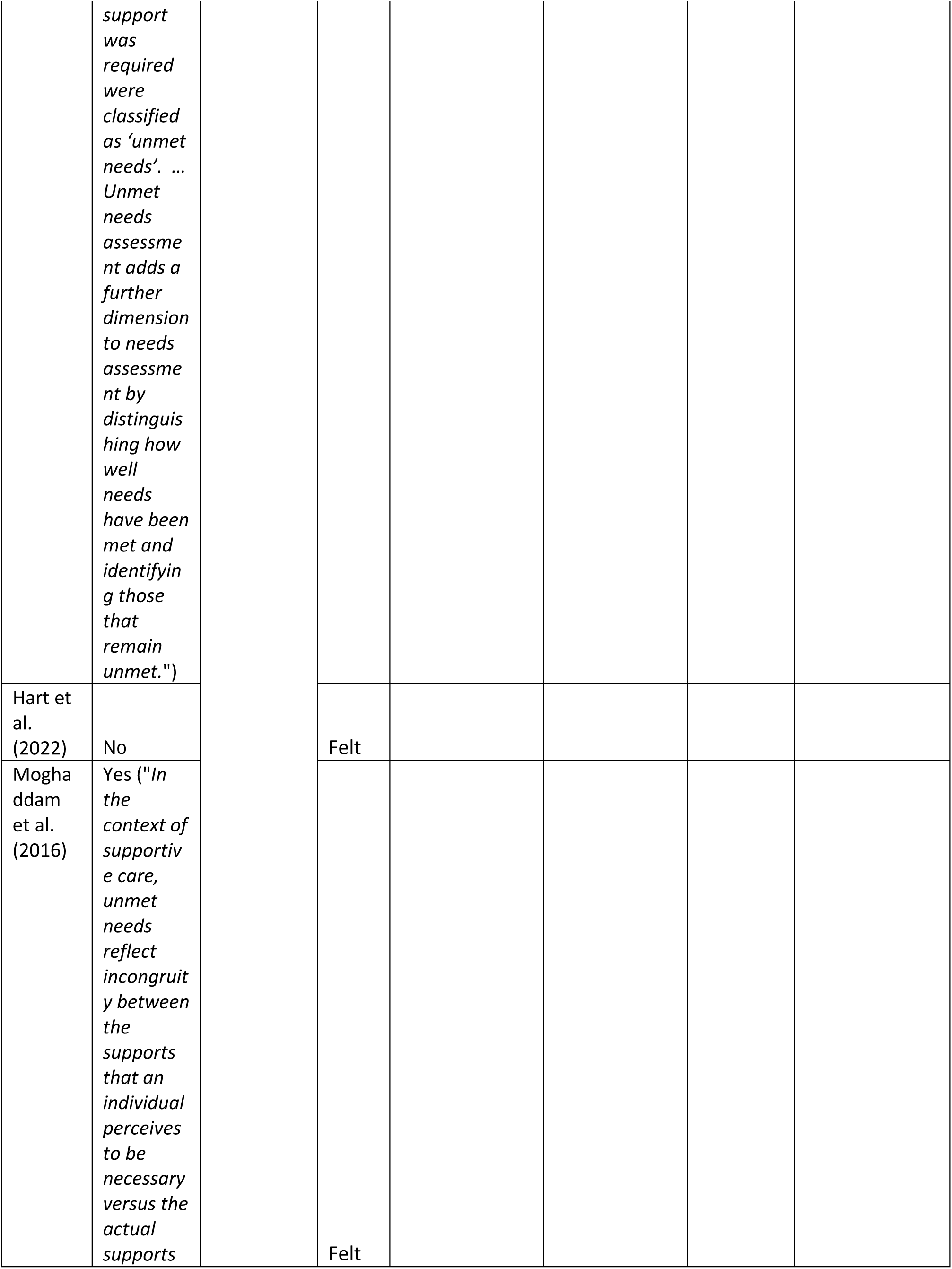
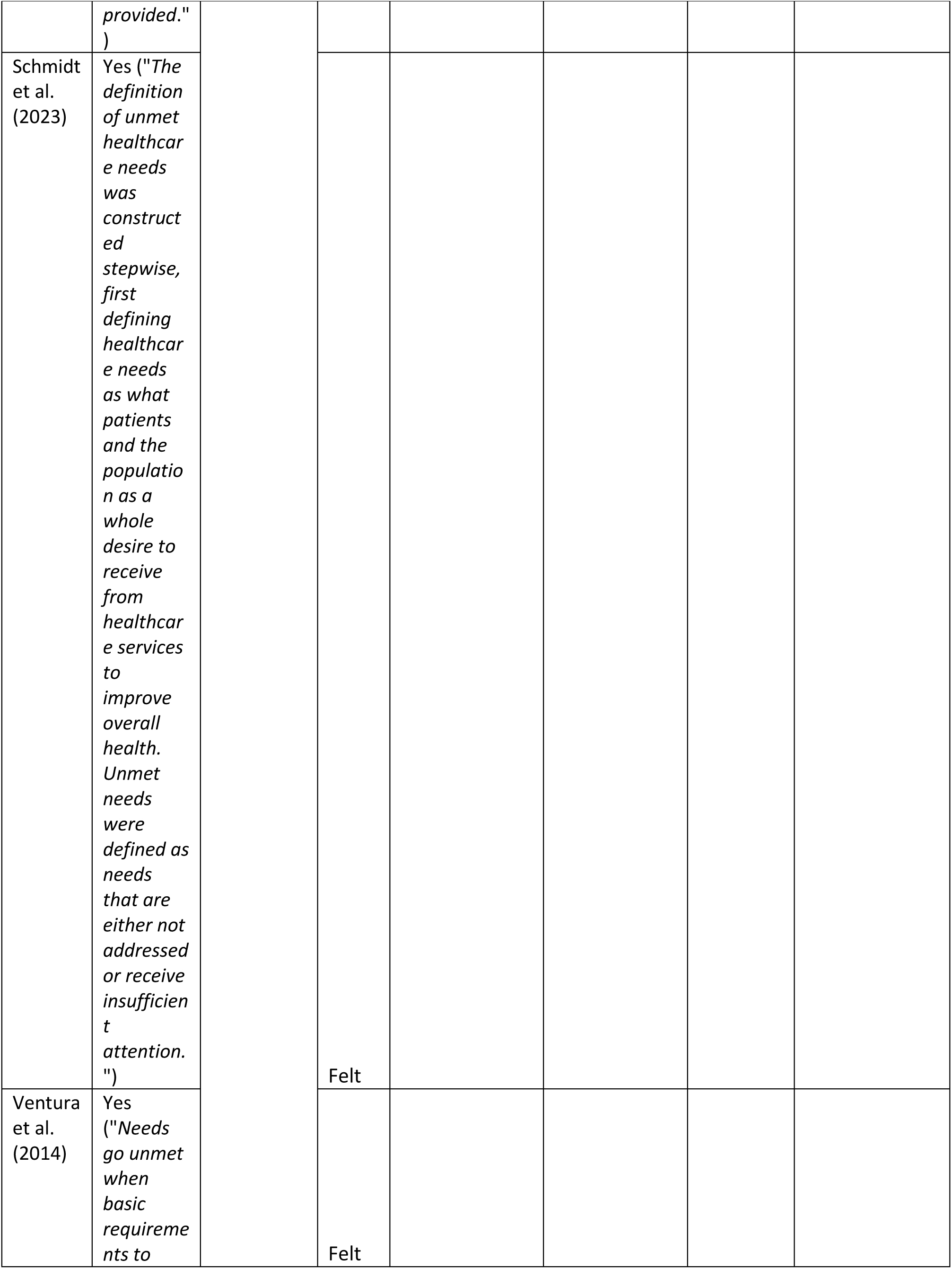
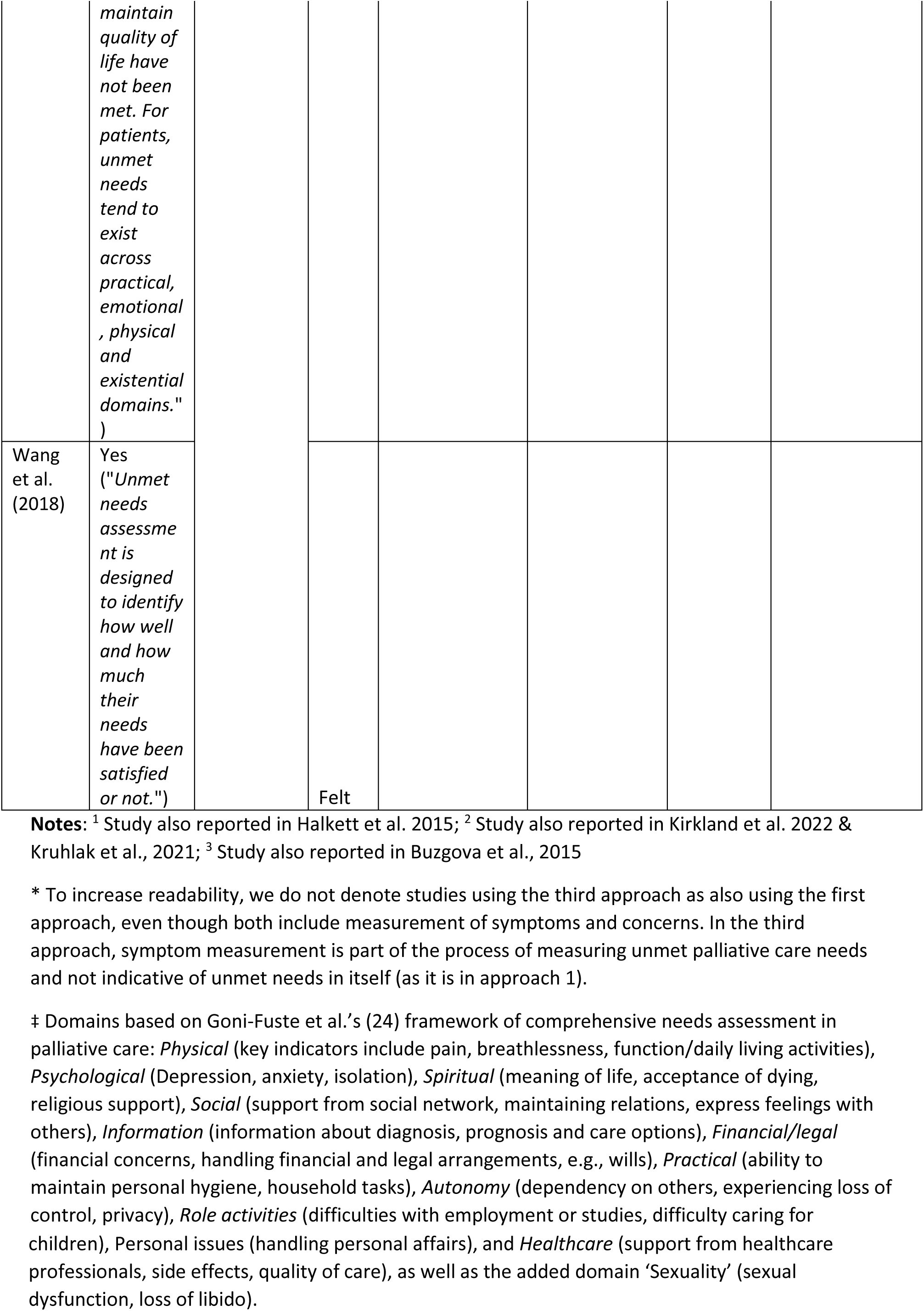
Definitions, measurement and reporting of unmet palliative care needs in the included studies (n=70).

Definitions varied but often emphasised a perceived discrepancy between the care required or desired and the care received, such as “the disparity between the supports that an individual perceives as necessary and those that are actually provided” (93). Other definitions highlighted presence of unfulfilled care needs, e.g., “needs that are either not addressed or receive insufficient attention” (97). Some studies focused on specific outcomes., e.g., conceptualising unmet needs as lacking “the requirement of some action or resource that is necessary, desirable or useful *to attain optimal well-being*" (33) or “when basic requirements *to maintain quality of life* have not been met” (98).

Partial definitions were usually presented as part of the study rationale, to suggest that care needs in specific patient groups or care settings might not be met. One study stated that “problems in the quality of life are not always correctly identified, and needs for care sometimes remain unmet” (88). Other studies implied that known issues of under-utilisation of palliative care meant that care needs might not be met, such as arguing that “Deficits in [palliative] care in any domains may result in multidimensional, unmet needs" (43).

Where studies specified care services that can help address unmet needs in their definition, few were explicit about which these services were or made any distinction between specialist and generalist palliative care.

### Measurement of unmet palliative care needs

#### Approaches to measurement

While few studies defined unmet palliative care needs, most described the approach used to measure it (i.e., the operationalisation), whether explicitly or implicitly. We identified three main approaches to measuring unmet palliative care needs: 1) *Symptoms and concerns* (studies using symptom prevalence to indicate unmet needs, without explicitly determining whether these symptoms were addressed or not); 2) *Access to services* (studies focusing on measuring service referrals or contacts, with little consideration of care outcomes); and 3) *Sufficiency of service provision to resolve symptoms and concerns* (studies addressing a perceived gap between the care provided and care deemed necessary to resolve symptoms and concerns). These approaches were not mutually exclusive and occasionally combined to varying degrees. Examples of each approach are presented in Table 4.

**Table 4.** Examples of approaches to measuring unmet palliative care needs in the included studies.

| Approach to measurement | Example of approach as described in the included studies |
| --- | --- |
| **Symptoms and concerns** | Unresolved needs were defined as having [Integrated Palliative care Outcome Scale (IPOS)] symptom scores above 2 [being moderately affected by a symptom or concern].<br>Hamano et al., 2023 (47) |
| **Access to services** | Unmet need was estimated as the difference in the proportion of decedents who were estimated to have needed palliative care [according to ten pre-determined conditions] and those who accessed relevant services [admission to one of the designated palliative care units or an episode of care from a palliative care specialist in any other setting].<br>Westley-Wise et al., 2022 (87) |
| **Sufficiency of service provision to address symptoms and concerns** | Unmet need was indicated when the patient responded that they experienced a problem either very much or to some extent and would have found additional assistance very or somewhat helpful.<br>Hwang et al., 2004 (55)<br>A need was defined as unmet when the respondent reported that the patient did not receive any or not enough help with that symptom.<br>Teno et al., 2004 (79) |

##### 1. Symptoms and concerns

The first approach to measurement of unmet needs relates to identifying and reporting the presence of one or more symptoms or concerns. Descriptions of what constituted a symptom or concern varied. Some studies narrowly focused on certain levels of specific symptoms, e.g., prevalence of pain being operationalised as having unmet need for pain relief (34). Other studies acknowledged accumulation of several symptoms or concerns, or these being present across various domains as indicating unmet need (43, 47), while others included anything that the patient deemed a problem or concern (76). Studies focusing on symptoms and concerns were often based on self or proxy reports of felt needs (in contrast to normative needs).

##### 2. Access to services

The second approach to measurement of unmet needs relates to the access to or provision of care. This includes absence of referral to specific services (such as specialist palliative care) for people who might benefit from this care, e.g., due to having specific diagnoses or other clinical characteristics.

Access to palliative services (whether generalist or specialist) was generally presumed to meet needs and often described in terms of maintaining or improving quality of life, whereas lack of contact with palliative services was indicative of unmet needs. These studies were often larger-scale studies using medical records (73) or clinical screening tools, e.g., the Palliative Care and Rapid Emergency Screening (P-CaRES) tool (61), to make inferences about people missing out on palliative care receipt at cohort or population level. As such these largely represent normative needs, i.e., as determined by professionals (9). This approach seldom distinguished between types of palliative care services, e.g., studies reporting overall rates of palliative care referrals (40, 42, 77). Availability of palliative care services was generally assumed, with only one study explicitly considering the lack of services across geographical regions (75).

##### 3. Sufficiency of service provision to resolve symptoms and concerns

Most studies’ approaches to measuring unmet palliative care needs related to insufficient care provision. This included the presence of symptoms, concerns or problems that were either not addressed at all, not adequately addressed (according to the patient or their family), or not satisfactorily resolved (for which the patient wanted further professional attention). This approach emphasises a gap or discrepancy between a desired or expected standard of care and what has been provided. Studies often measured insufficient provision of care by asking patients or proxies (usually patients’ families or, less commonly, healthcare professionals) first whether a problem or concern was present and then the extent to which it was adequately provided for and addressed, thus primarily capturing felt need (9). This approach was primarily used by small-scale survey studies using convenience samples to describe patients’ perspectives on the presence of unresolved issues, although some were larger scale (51, 79).

##### Sources of information for measuring unmet palliative care needs in primary evidence studies

Among primary evidence studies that measured unmet need, methods for measurement varied in terms of who and what was the source of information for the assessment (see Table 3). Most studies used self-reported data (n=39, 68%) collected using research questionnaires or patient reported or patient-centred outcome measures (PROMs or PCOMs). Proxy-reported data by bereaved family members, e.g., using mortality follow-back surveys were also common (n=15, 25%). These methods largely capture felt needs (i.e., an individual’s perceived need for care). Clinical assessments by healthcare professionals (often through use of screening tools) (n=8, 11%) or researchers (n=8, 11%) were less common, representing normative need (9).

##### Instruments and timing of measurement in primary evidence studies

As shown in Table 3, named instruments or measures were used in 42 (69%) of the primary evidence studies, whereas nine (15%) used unnamed questionnaires, and one (2%) used an unnamed structured interview. The most used named measure was the Supportive Care Needs Survey - Short Form (SCNS-SF34) (n=8, 13%), followed by the Problems and Needs in Palliative Care - short version (PNPC-sv) (n=5, 8%), both of which were initially developed for cancer patients. Unmet needs were estimated using administrative medical records in nine (15%) studies.

Almost half did not report the time period during which unmet needs were measured (n=30, 49%). Of those that specified a time interval, self-reported unmet needs commonly related to the past month (n=15, 25%). In studies using samples of decedents, the time interval of interest was usually the last month of life (n=4, 7%) or last week of life (n=3, 5%). Only three studies reported longitudinal measurements of unmet needs over time (47, 83, 89).

##### Reporting of unmet palliative care needs in primary evidence studies

Some of the 61 primary evidence studies reported unmet needs at the item-level, usually as the proportion of patients with a specific unresolved symptom (34, 39, 80). Most, however, reported unmet needs at the domain-level by combining responses to several items related to a specific domain of care (e.g., (65, 69)). Our mapping of the unmet care needs reported in the primary evidence captured eleven distinct domains of needs (Table 3). Ten of these were identified in the framework by Goni-Fuste et al. (2021), with the addition of “Sexuality” which was reported in 14 (23%) studies. Of the 52 primary evidence studies that measured unmet palliative needs using a questionnaire or interview, most used multi-domain measures that allowed a holistic assessment of unmet palliative care needs: 38 (73%) captured three dimensions or more, while only four (8%) focused solely on one domain (spiritual needs (48, 66, 69)) and physical function (56)).

In total, 26 studies (43%) reported one single metric of unmet palliative care needs, usually as a percentage of participants needing palliative care but not adequately receiving it. These rates ranged from 10% in a 2023 study of hospitalised patients in United States (73) to 30% among people with non-cancer conditions in a 2008 health omnibus survey in Australia (42). Other studies calculated unmet needs by aggregating items and/or domains from the measure, which was commonly reported as prevalence of having one or more unmet needs (e.g., “71.8% of patients reported at least one unmet supportive care need” (83)), sometimes with reference to severity (e.g., “48% reported one or more moderate or high unmet needs” (86)). Some studies presented a mean or median number of unmet needs within the group (e.g., “The median number of total unmet needs identified was 3” (74)).

## Discussion

### Key findings

This scoping review examines how existing evidence has defined, measured and reported unmet palliative care needs across disease groups and care settings. Only 11 out of 70 (16%) defined the theoretical construct of unmet needs. The review also found variability in approaches to measurement and reporting across studies, identifying three main approaches to measuring unmet palliative care needs: 1) the presence or severity of symptoms and concerns; 2) the level of access to palliative care services; and 3) symptoms and concerns that remain unresolved despite service provision.

Unmet palliative care needs were commonly identified using self-reported data about the presence of symptoms and concerns and whether these were resolved by the care provided. Similar to previous reviews on unmet palliative care needs in the emergency department (100) and in nursing homes (101), we found that the lack of explicit definition hinders consistent measurement and comparisons across studies, making it difficult to determine how best to measure or estimate unmet need at the population-level. This lack of clarity creates ambiguity about what is being assessed and risks conflating unmet needs with related concepts, such as satisfaction with care or quality of life. There is also a conceptual distinction between unmet needs as outcomes (e.g. inadequate pain relief), and reasons for unmet needs (e.g., not accessing to palliative care services), which was rarely noted in the included studies.

Expanding on the findings of Franks et al. (12) on population-level palliative care needs assessments, our review identifies three approaches to measuring unmet needs in the existing literature. The first approach, *Symptoms and concerns*, involves quantifying prevalence or burden of problems. This approach corresponds to Franks et al. (12) ‘epidemiological approach’ and is not dependent on whether patients are known to services or not. Patient reported or patient-centred outcome measures (PROMs and PCOMs) can be very helpful for collecting these data and, depending on how many domains a measure captures, this approach enables a holistic assessment of palliative care needs. For example, the Integrated Palliative care Outcome Scale (IPOS) is a valid and reliable measure for multi-domain symptoms and concerns, allowing evaluation of the complexity of needs for care (102). Wide international use of PROMs and increasing integration with patient record systems further aids access to comparable data for research. Nevertheless, this approach has limitations, as having unmet needs do not necessarily mean professional help is desired, and it may not clarify which professionals or services should meet any needs identified.

The second approach, *Access to services*, infers unmet palliative care needs from access to services, aligning with Franks et al.’s (12) healthcare utilisation approach. Service use is usually documented in medical records and often readily available in national patient registers, which facilitates their use in research, policymaking, and commissioning. While evaluating access to services, e.g., referrals, can elucidate care gaps or inequities indicative of underserved populations, this approach requires that patients are correctly identified as having palliative care needs in the first place (103). Moreover, uptake of palliative care services might not reflect actual needs for care (42), and access to services alone cannot indicate whether needs are met or if the care delivered improves the patient’s situation (90).

The third approach identified in this review, *Sufficiency of service provision to resolve symptoms and concerns*, is the most comprehensive as it combines aspects of the first two in operationalising unmet needs (presence of symptoms and concerns but also whether services adequately addressed these). Studies using this approach more often distinguished between having an unmet problem and desire for care, showing that patients did not necessarily want professional attention even for serious problems, especially within social, sexual and spiritual domains (57, 69, 88). This approach is helpful for evaluating outcomes of care and person-centredness (104), i.e., not only determining whether symptoms are present, or care delivered, but whether the care met individual needs.

Focusing on the gap between desired and received care can risk overestimating unmet needs if expectations of symptom relief at the end of life are unrealistic. For example, a symptom like fatigue (105) is known to be common towards the end-of-life but difficult to fully relieve even with optimal currently available treatment. Studies adopting this approach must consider the subjective nature of measurement and how the person making the assessment might influence estimations of unmet need, since reports of symptom relief can vary between family members and healthcare professionals (106).

Each approach has its strengths and limitations, and all can help estimating unmet palliative care needs, depending on context and research question. For example, the *Symptoms and concerns* approach is well suited for intervention studies to assess changes in needs over time in response to a care intervention. The *Access to services* approach is helpful for service mapping studies, and the *Sufficiency of service provision to resolve symptoms and concerns* approach could be used to inform future post-bereavement survey research. Appropriateness of the approaches may also vary according to the healthcare system from which the evidence is derived (107). As already mentioned, the approaches do not distinguish between which services should meet which level of unmet needs. Future research should examine how variation in service use might be associated with management of needs across the physical, psychological, spiritual and social domains. The high proportions of unmet needs reported in the included studies also raise questions about ‘acceptable’ levels of unmet needs at the population-level. The definitions of unmet needs presented in this review covered a wide span, ranging from care for optimal well-being (33) to basic requirements for maintaining quality of life (98), and provide little clarity about what constitutes a feasible standard.

### What this review adds

Resource limitations invariably mean that specialist palliative care access is prioritised for the most urgent and complex cases (108), and it is not surprising that the existing research has focused on identifying specialist palliative care needs. However, better understanding of whether specialist palliative care services meet patients’ needs is crucial to inform decision-making about funding, planning and delivering high-quality care (109). Despite 25 years since Franks et al.’s (12) review highlighted poor-quality and inconsistent population-level estimates of palliative care needs, the literature is largely based on cross-sectional studies using self-reported data from convenience samples, with few studies using population-based or representative samples.

To provide a comprehensive understanding of unmet palliative care needs, multi-domain multi-level assessments like those presented in the third approach to measurement are well-suited to capture both felt and normative need, particularly when combining objective data sources, like service provision, and subjective experiences of the care received. In Box 1 we provide further recommendations for advancing research, policy and practice and building a more robust evidence-base for estimating the prevalence of unmet needs in the general population. Additional considerations of data collection methods are provided in Supplement file 3.

##### Box 1. General and audience-specific recommendations based on the review findings.

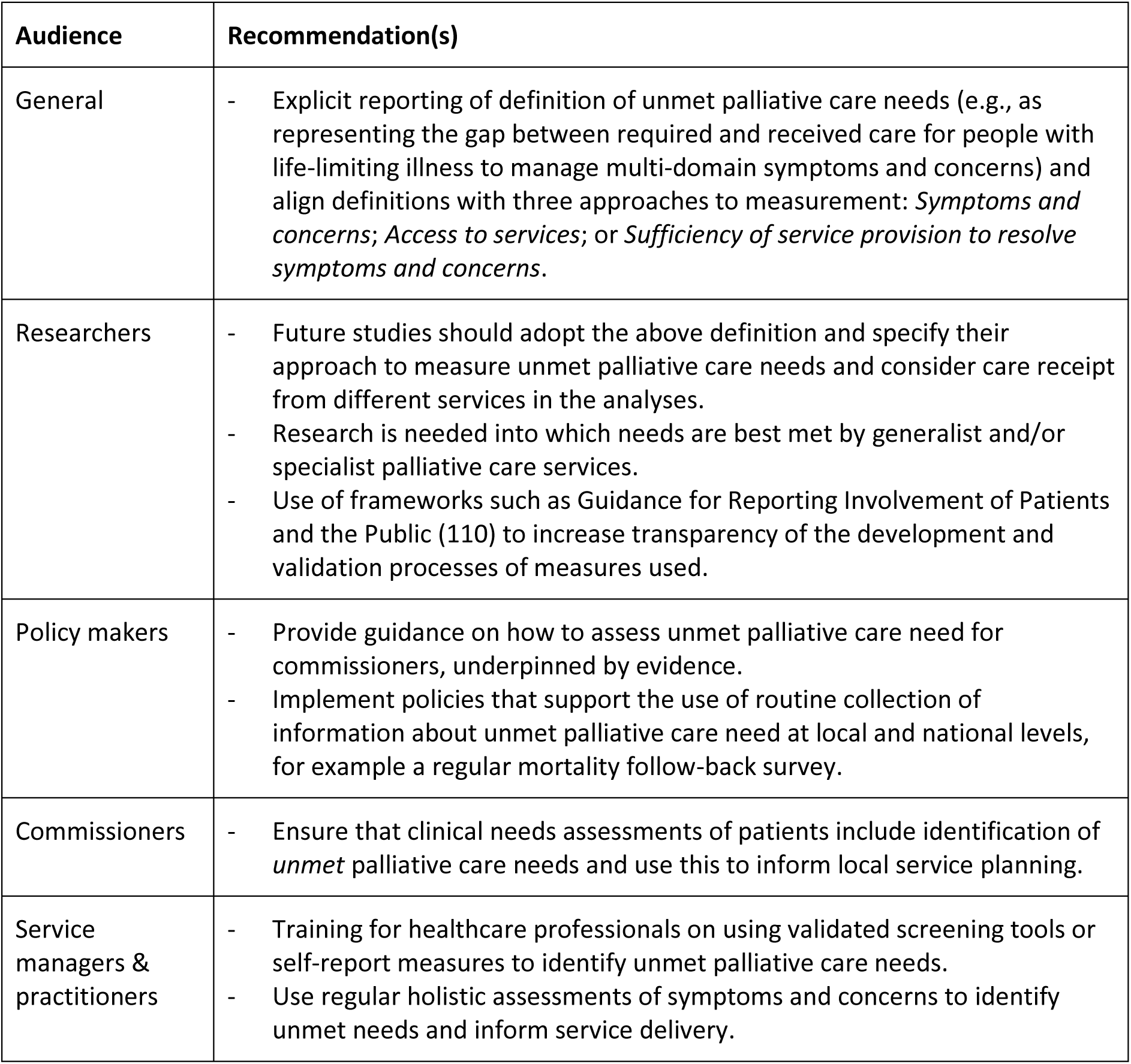

Routine data collection from continuous multi-domain needs assessments is crucial to advance the understanding of unmet palliative care needs at the population-level and inform policymakers and commissioners in the planning and delivery of palliative and end-of-life care (18, 111). For studies using administrative data from medical records, standardising the use of clinical variables in screening tools in care practice would enhance consistency in identification of unmet palliative care needs. Challenges remain, however, regarding how needs are disclosed, as limited awareness of available services or resources can hinder people from recognising unmet needs (112), and lack of trust and power imbalances can hinder requests for support, particularly for potentially sensitive topics such as mental health issues, living situations, or family relationships (74).

### Strengths and limitations

This is the first scoping review to summarise how unmet palliative care needs has been defined, measured and reported in existing literature. While prior reviews have focused on the psychometric properties of measures of unmet palliative care needs (13, 14, 113), this review contributes with new knowledge about overarching theoretical and operational aspects of the existing evidence on unmet palliative care needs.

A strength of this review is the inclusion of grey literature as well as use of forward and backward citation searches to mitigate the risk of omitting relevant sources. We also present data at the study-level, thereby avoiding double counting duplicate data sets across sources (24). Although the methodological variation of included studies hindered conclusions to be drawn regarding reported prevalence of unmet palliative care needs, this is not a limitation for the current study since scoping reviews aim to identify and describe the breadth of existing evidence and not synthesise study findings (114).

By not limiting our inclusion criteria to specific disease groups or care settings, we provide a comprehensive overview of how unmet palliative care needs has been researched in various contexts, with the objective to expand on the work of Franks et al. (12) and further the understanding of prevalence of unmet palliative needs are measured. However, our review also has limitations. Including only quantitative or mixed-methods studies and reviews reduces the range of evidence sources and may limits the completeness of the findings, as we might have missed important conceptual work from qualitative sources. The lack of quality appraisal and risk of bias assessment is another inherent limitation to scoping reviews (115) but also allows for a more rapid review process.

## Conclusion

This is the first scoping review to summarise the existing literature on how unmet palliative care needs have been defined, measured and reported across disease groups and care settings. Even though quantifying unmet palliative care needs is crucial for planning and equitable delivery of palliative care services, our review demonstrates that studies rarely explicitly define the construct and there is considerable variation in how unmet needs are measured and reported. Clear reporting of definitions and operationalisations of unmet palliative care needs is required to harmonise the evidence base and subsequently enable estimation of the proportion of people with unmet needs at the population-level. We identify three distinct approaches to measurement of unmet palliative care needs and provide guidance on their use for various end-users.

## Data Availability

This review is registered on the Open Science Framework (https://doi.org/10.17605/OSF.IO/M8DHA). The full search strategy is provided in the Supplementary material. Inclusion/exclusion criteria are presented in the article. Additional data will be made available upon request.

## Declarations

### Authorship

T.J. and M.D. were responsible for the design of the study, with critical input from all authors. M.D. and T.J. developed the search strategy, M.D. conducted the searches. T.J., M.D., A.F, and L.F. screened articles for inclusion. T.J., A.B., A.F., and M.D extracted data. T.J. charted the data; A.B., T.J and M.D. conducted qualitative synthesis. All authors contributed to analysis and interpretation of data. T.J. drafted and revised the manuscript. All authors critically revised the manuscript for intellectual content, and read and approved the final manuscript.

### Ethics and consent

Ethical approval was not required for this review as there was no direct patient contact or access to individual participant data.

