## Supplementary tables 1-3 for "Defining and measuring unmet palliative care needs among people with life-limiting illness: a scoping review of international evidence"

### Supplementary material

**Supplement file 1**. Search Strategy

Example Search Strategy for MEDLINE (Ovid) designed together with an Information Specialist at Kings College London (KCL) (search #1 adapted from the CareSearch Palliative Care search filter for PubMed).

| | |
|---|---|
| #1 | SH advance care planning OR SH attitude to death OR SH bereavement OR SH bereavement support SH terminal care OR SH hospice OR SH palliative therapy OR SH terminally ill patient OR SH death OR palliat* OR hospice* OR terminal care OR advance care plan* OR attitude to death OR bereavement OR terminal care OR life supportive care OR terminally ill |
| #2 | SH Unmet Medical Need OR unmet adj3 (need* OR demand*) OR for?go OR for?gone |
| #3 | #1 AND #2 |

**Supplement file 2:** Data Extraction Form

| **Data extraction category** |
|---|
| 1. Authors |
| 2. Year of Publication |
| 3. Country |
| 4. Type of Evidence Source |
| 5. Aim and Objectives |
| 6. Study Design |
| 7. Study period |
| 8. Population |
| 9. Setting |
| 10. Sampling technique |
| 11. Representativeness of Sample |
| 12. Sample Size |
| 13. Methods |
| 14. Type of Care |
| 15. Care need definition |
| 16. Unmet care need definition |
| 17. How have unmet needs been operationalised? |
| 18. Measurement |
| 19. Domains of unmet need measured? |
| 20. Who assesses the unmet need? |
| 21. Reported measurement results |
| 22. Key findings or conclusions relating to the review question |
| 23. Forward citation search result (Scopus) |

**Supplement file 3**. Considerations of different data collection methods to estimate unmet palliative care needs across the three approaches measurement identified in the review.

| Method | Measurement approach(es) | Strength(s) | Limitation(s) |
|---|---|---|---|
| Patient reported/centred outcome measures (PROMS/PCOMS) | • Symptoms and concerns<br>• Sufficiency of service provision to resolve symptoms and concerns | • Directly reported by patients, thereby increasing validity<br>• Allows for more complete or nuanced understanding of needs<br>• Services can respond to any unmet palliative care needs identified and signpost patients and families for further support | • Can be a burdensome task for a vulnerable patient group<br>• Patients may not be able to complete due to declining health<br>• Patients may not have complete information about available services and what they should expect from them |
| Clinical needs assessment by health professionals | • Symptoms and concerns | • Provides good knowledge of what needs can be addressed by services<br>• Potential to be more complete than patient reported measures | • May not fully reflect the felt needs of the patient, e.g., due to not feeling comfortable disclosing certain problems. |
| Administrative medical records | • Access to services | • Efficient way of estimating unmet need in large populations using conventional proxies for unmet needs. | • Currently largely reliant on service utilisation and other crude indicators of care needs such as diagnosis, and therefore unlikely to be informative about patients' subjective experiences of unmet needs |
| Mortality follow-back survey by family members | • Symptoms and concerns<br>• Access to services<br>• Sufficiency of service provision to resolve symptoms and concerns | • Families' experiences of their relative's final months of life can provide useful and detailed information about how well palliative care needs were met.<br>• Recurring measurement can be used to evaluate how well services are responding to needs over time<br>• Focuses on deaths during a defined time-period, and allows comparison across patient groups, areas etc | • Recall bias may limit the validity<br>• Family members may not have complete information about what services were accessed, or which services are available and what care they provide.<br>• Family members may overstate needs or not have realistic expectations of care towards the end of life. |
| Population based health surveys, e.g., omnibus surveys | • Symptoms and concerns<br>• Access to services | • Efficient way of reaching many people, including patients and family carers. | • Less precise way of attaining information about peoples care in the last few months of life, as the survey |

| | | | |
|---|---|---|---|
| | • Sufficiency of service provision to resolve symptoms and concerns | • Data collection can be integrated with other health-related research projects, possible reaching people who might not want to participate in an end-of-life care focused survey | is not targeted at those with recent experience. |
